# Multi-omics analysis reveals the impact of microbiota on host metabolism in hepatic steatosis

**DOI:** 10.1101/2021.05.22.21257482

**Authors:** Mujdat Zeybel, Muhammad Arif, Xiangyu Li, Ozlem Altay, Mengnan Shi, Murat Akyildiz, Burcin Saglam, Mehmet Gokhan Gonenli, Buket Yigit, Burge Ulukan, Dilek Ural, Saeed Shoaie, Hasan Turkez, Jens Nielsen, Cheng Zhang, Mathias Uhlén, Jan Borén, Adil Mardinoglu

## Abstract

Non-alcoholic fatty liver disease (NAFLD) is a complex disease involving alterations in multiple biological processes regulated by the interactions between obesity, genetic background and environmental factors including the microbiome. To decipher hepatic steatosis (HS) pathogenesis by excluding critical confounding factors including genetic variants, obesity and diabetes, we characterized 56 heterogeneous NAFLD patients by generating multi-omics data including oral and gut metagenomics as well as plasma metabolomics and inflammatory proteomics data. We explored the dysbiosis in the oral and gut microbiome and revealed host-microbiome interactions based on global metabolic and inflammatory processes. We integrated this multi-omics data using the biological network and identified HS’s key features using multi-omics data. We finally predicted HS using these key features and validated our findings in a validation dataset, where we characterized 22 subjects with varying degree of HS.

**Significance statement:** The oral and gut microbiota alterations have been linked to NAFLD. There is a lack of data on multi-omics characteristics of hepatic steatosis by exclusion of major confounding factors of obesity and metabolic syndrome. We observed that the oral and gut microbiota remodelling starts at early stages of the NAFLD spectrum, independent of obesity and metabolic syndrome. Our analysis suggested that the bacterial diversity is correlated with multi-omics signatures in NAFLD and our predictive model created based on multi-omics variables can successfully predict hepatic steatosis. The components of the multi-omics signatures may serve as biomarkers and can be pharmaceutically targeted. Future clinical trials with microbiota manipulation could consider intervention at early stages of NAFLD.

## Introduction

Non-alcoholic fatty liver disease (NAFLD) is characterized by deposition of lipid droplets in the liver without significant alcohol consumption and secondary causes. NAFLD constitutes a wide range of the clinical spectrum, including hepatic steatosis (HS) and non-alcoholic steatohepatitis (NASH), which may ultimately lead to advanced fibrosis and cirrhosis. In parallel to ongoing epidemics of obesity, NAFLD incidence is increasing globally, affecting almost one-fourth of the population (Abeysekera *et al*, 2020; Bugianesi, 2020). Even though NAFLD is becoming a leading aetiology of chronic liver disease, therapeutic approaches for NAFLD are currently limited with lifestyle modifications encompassing dietary intervention and physical activity (Alferink *et al*, 2019; Romero-Gómez *et al*, 2017; Sanyal, 2019)

Given that NAFLD is closely linked to metabolic syndrome, obesity and type 2 diabetes (T2D), its pathogenesis is remarkably complex. Discoveries of *PNPLA3* and *TM6SF2* variants have highlighted the pathological processes leading to metabolic disturbances in HS. However, only 10-20% of NAFLD susceptibility could be attributed to known genetic variants (Eslam & George, 2020). Exploring the connections between the hepatic and extra-hepatic metabolic factors through the generation of the multi-omics data may reveal the underlying molecular mechanisms associated with the disease’s occurrence, discovering novel biomarkers and drug targets, and eventually provide further insight for the development of efficient treatment strategies. The dysbiosis and diversity in the oral and gut microbiome have been associated with NAFLD (Boursier *et al*, 2016; Mardinoglu *et al*, 2018c) as well as metabolic syndrome (Vrieze *et al*, 2012; Yuan *et al*, 2019), type 2 diabetes (Mardinoglu *et al*, 2019) and obesity (Leung *et al*, 2016). Studies indicated that microbiome composition changes are associated with advanced hepatic fibrosis (Loomba *et al*, 2017) and cirrhosis (Bajaj *et al*, 2014; Lelouvier *et al*, 2016). Moreover, several multi-omics studies integrated complementary –omics data across various environmental states through systems biology. These studies demonstrated that a multi-omics approach is a powerful tool for understanding the dynamics of biological functions in liver diseases and other associated metabolic conditions (Altay *et al*, 2019; Mardinoglu *et al*, 2018a).

Here, we performed an in-depth characterization of 56 subjects to decipher NAFLD pathogenesis by excluding critical confounding factors including obesity, diabetes and genetic variant-associated HS (Figure 1A). We collected saliva and faeces samples for studying the dysbiosis in the oral and gut microbiome through the generation of shotgun metagenomics data and identified the key species involved in various stages of HS. Moreover, we performed plasma metabolomics and inflammatory proteomics analysis to explore the host-microbiome interactions. We studied the altered global metabolic and inflammatory processes, and its connections with the species’ abundances in the oral and gut microbiome. We integrated this multi-omics data using biological networks and identified key features of HS using multi-omics data. We finally predicted HS using these key features and validated our findings in an independent validation dataset, where we characterized 22 subjects with varying HS.

**Figure 1.**
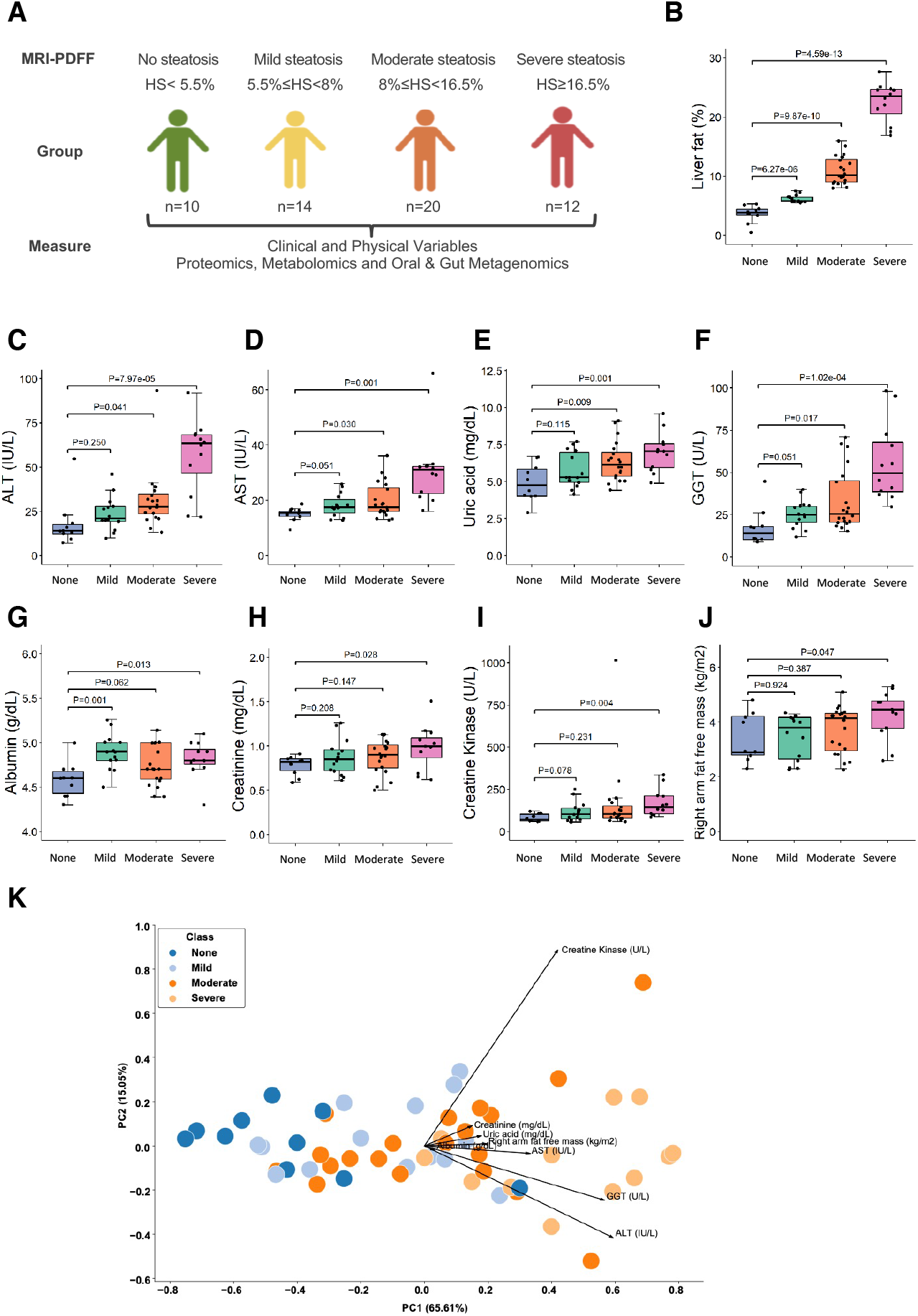
(A) The design of the study. Subjects were stratified into four distinct groups based on the hepatic steatosis percentages measured using MRI-PDFF. (B) The boxplot shows hepatic steatosis of subjects with none, mild, moderate and severe steatosis. (C-J) Significantly different clinical parameters are presented in subjects with none, mild, moderate and severe steatosis (K) PCA of all subjects based on eight significantly different clinical and physical variables shows good separation.

## Results

### The associations between HS and clinical and physical variables

We generated data for 56 overweight and obese subjects (BMI>28.8) with a varying degree of HS. We excluded patients if they carry PNPLA3 I148M (homozygous for I148M). We determined HS by the proton density fat fraction, measured by magnetic resonance imaging (MRI-PDFF). We classified the subjects into four different groups: i) 10 subjects with no steatosis (HS<5.5%), ii) 14 subjects with mild steatosis (5.5%≤HS<8%), ii) 20 subjects with moderate steatosis (8.0%≤HS<16.5%) and iv) 12 subjects with severe steatosis (HS ≥16.5%) (Figure 1A-B). We carefully phenotyped these subjects by measuring clinical and physical variables (Dataset S1A-C). We also collected saliva and faeces samples to generate metagenomics data, and plasma samples to generate metabolomics and inflammatory proteomics data. The subjects’ demographic characteristics in each group are presented (Dataset S1C-D).

To confirm our findings based on multi-omics analysis of 56 subjects and avoid the genetic differences between the subjects, we re-analysed 22 of these patients after 2-3 months, generated clinical data and measured the HS. We observed that the degree of HS in each subject has changed since they have been recommended changes in their exercise and eating habits. The subjects’ demographic characteristics in each group are presented (Dataset S2A-D). These 22 subjects were classified based on HS as i) two subjects with no steatosis, ii) five subjects with mild steatosis, iii) eight subjects with moderate steatosis, and iv) seven subjects with severe steatosis. In the validation dataset, we also generated oral (Dataset S2E) and gut (Dataset S2F) metagenomics, metabolomics (Dataset S2G) and proteomics (Dataset S2H) data using a similar methodology as in the finding dataset.

We analyzed the differences in the clinical and physical variables between the groups with different HS degrees in the finding dataset (Dataset S1C). We did not find any differences in the weight, BMI, waist circumference, HOMA-IR score, glucose, insulin and HbA1c levels between different steatosis groups. We found that the levels of alanine aminotransferase (ALT) (Figure 1C), aspartate aminotransferase (AST) (Figure 1D), uric acid (urate) (Figure 1E) and GGT (Figure 1F) were significantly higher in subjects with severe and moderate steatosis but not mild steatosis, compared with no steatosis. Although we could not detect any significant differences in the level of these different clinical parameters in subjects with mild steatosis vs no steatosis, we observed a tendency of decrease in these variables’ grades. We also found that the levels of clinical variables including albumin (Figure 1G), creatinine (Figure 1H) and creatine kinase (Figure 1I) as well as the amount of fat-free mass in right arm (Figure 1J) were significantly higher in subjects with severe steatosis but not in subjects with moderate and mild steatosis, compared with no steatosis. Similarly, we observed a tendency of decrease in clinical and physical variables in subjects with moderate and mild steatosis vs no steatosis. Based on these variables, we performed Principal Component Analysis, and this showed a distinct separation between no, mild, moderate and severe steatosis (Figure 1K).

### Dysbiosis in the gut and oral microbiome of NAFLD patients

We generated metagenomics data to study the dysbiosis in the microbial composition in the gut and oral microbiome (Dataset S3) and explored the interactions between the host and microbiome in subjects with varying degree of HS. We first compared the differences in the species’ abundances between mild vs no steatosis in the gut microbiome. We found that the abundances of individual species in Bacteroidetes (*Prevotella* sp CAG 520, *Prevotella* sp AM42 24, *Butyricimonas virosa* and *Odoribacter splanchnicus*), Proteobacteria (*Escherichia coli*), Lentisphaerae (*Victivallis vadensis*) and Firmicutes (*Holdemanella biformis, Dorea longicatena, Allisonella histaminiformans* and *Blautia obeum*) were significantly reduced in subjects with mild steatosis vs no steatosis. Notably, when we compared moderate vs no steatosis, we found that the abundance of only *Firmicutes bacterium* CAG 95 was significantly reduced. In contrast, the abundance of species belongs to Firmicutes (*Streptococcus mitis* and *Roseburia inulinivorans*), and Bacteroidetes (*Barnesiella intestinihominis* and *Bacteroides uniformis*) were significantly increased in subjects with moderate steatosis vs no steatosis (P<0.05, Figure 2A, Dataset S4). Moreover, we compared the species’ abundances in the gut microbiome between severe vs no steatosis patients. We found that the abundances of the species in Actinobacteria (*Slackia isoflavoniconvertens*), Bacteroidetes (*Butyricimonas virosa* and *Odoribacter splanchnicus*), Lentisphaerae (*Victivallis vadensis*), Firmicutes (*Dorea longicatena, Firmicutes bacterium* CAG 83, *Firmicutes bacterium CAG 95, Firmicutes bacterium* CAG 110, *Roseburia hominis, Roseburia sp* CAG 182, *Oscillibacter sp* CAG 241 and *Ruminococcus bromii*) and Proteobacteria (*Bilophila wadsworthia*) were significantly reduced in subjects with severe steatosis vs no steatosis (P<0.05, Figure 2A, Dataset S4). We observed that the abundance of *Firmicutes bacterium CAG 95* was significantly reduced in the gut microbiome of subjects with both severe and moderate steatosis vs no steatosis.

**Figure 2.**
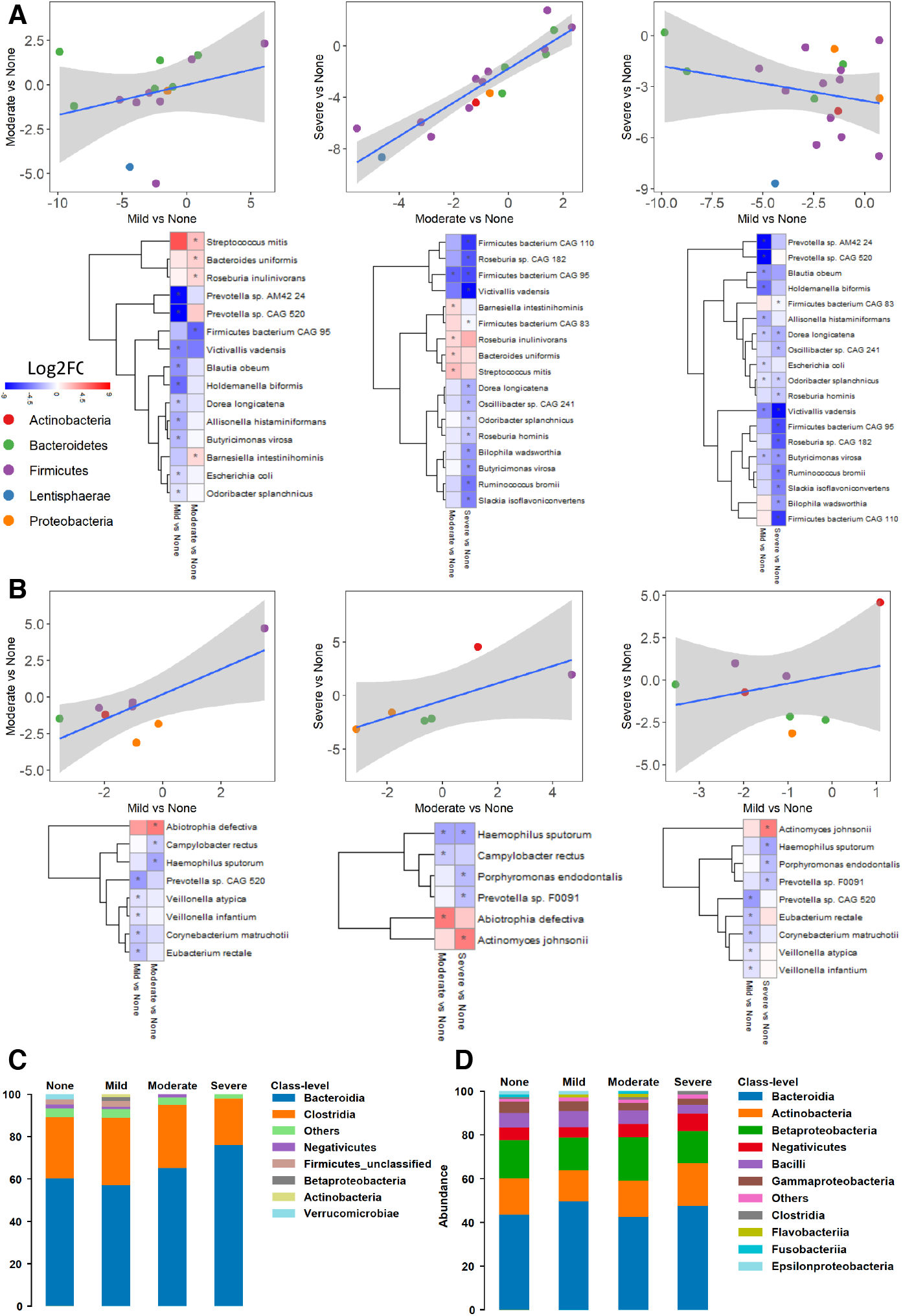
A) Scatter plot with a linear regression line, and heatmap shows Log2FC based alterations of the significantly different species in the A) gut microbiome and B) oral microbiome of subjects with different hepatic steatosis degrees. Asterisks indicate statistical significance based on paired Wilcoxon signed-rank tests. P<0.05. Log2FC: log2(fold change). Stacked bar plots depicting class-level differences in C) gut microbiome and D) oral microbiome composition between the severe-no, moderate-no and mild-no steatosis groups. The “other” subcategory included viruses, fungi, and rare species (abundance <1%).

Similarly, we compared the differences in the species’ abundances between mild vs no steatosis in the oral microbiome. We found that the abundance of the specific species in Firmicutes (*Veillonella atypica, Veillonella infantium* and *Eubacterium rectale)*, Bacteroidetes (*Prevotella* sp CAG 520) and Actinobacteria (*Corynebacterium matruchotii*) were significantly reduced in subjects with mild steatosis vs no steatosis (P<0.05, Figure 2B, Dataset S4). Our findings also revealed that increased abundance of species in Firmicutes (*Abiotrophia defectiva*) and reduced abundance of Proteobacteria (*Campylobacter rectus* and *Haemophilus sputorum)* in subjects with moderate vs no steatosis (P<0.05, Figure 2B, Dataset S4). Notably, the abundance of species in Bacteroidetes (*Porphyromonas endodontalis* and *Prevotella* sp F0091) and Proteobacteria (*Haemophilus sputorum*) were significantly reduced, whereas the abundance of species in Actinobacteria (*Actinomyces johnsonii*) were significantly increased in the oral microbiome of subjects with severe steatosis vs no steatosis (P<0.05, Figure 2B, Dataset S4). We observed that the abundance of *Haemophilus sputorum* was significantly reduced in the oral microbiome of subjects with both severe and moderate steatosis vs no steatosis.

In line with the severity of steatosis, we found that Bacteroidia was the most, and Clostridia was the second most abundant bacteria in the gut microbiome composition. The Clostridia/Bacteroidia ratio is notably decreased in severe steatosis vs no steatosis (Figure 2C, Dataset S3). Moreover, we found that the relative abundance of the Firmicutes and Negativicutes is reduced in severe steatosis vs no steatosis. On the other hand, there was more diversity in the oral microbiome than the gut of subjects with steatosis; where the abundance of the Negativicutes was increased in severe steatosis vs no steatosis (Figure 2D, Dataset S3).

### The associations between metagenomics data and clinical parameters

Correlation analysis between the abundances of oral and gut microbial species and the significantly altered clinical variables showed that there are 2 distinct clusters with the reduced and increased abundance of species (Figure 3A). Our analysis showed that the abundance of species belongs to Firmicutes (*Ruminococcus bromii*) and Bacteroides (*Sanguibacteroides justesenii*) in the gut was negatively correlated with all HS, AST, ALT, GGT and uric acid levels (P<0.05, Figure 3A, Dataset S5). Additionally, we found that the abundance of species belongs to Firmicutes (*Dorea longicatena* and *Roseburia* sp CAG 182) was negatively correlated with HS, AST, ALT and uric acid levels (P<0.05, Figure 3A, Dataset S5). In an extended manner, we observed that the reduced abundance of individual species in Bacteroides (*Sanguibacteroides justesenii*), Firmicutes (*Firmicutes bacterium* CAG 95, *Firmicutes bacterium* CAG 110, Firmicutes bacterium CAG 238, *Lactobacillus ruminis, Phascolarctobacterium* sp CAG 266), Lentisphaerae (*Victivallis vadensis*) and Proteobacteria (*Bilophila wadsworthia*), as well as the increased abundance of species, belongs to Proteobacteria (*Haemophilus* sp HMSC71H05) in the gut were significantly correlated with the HS (P<0.05, Figure 3A, Dataset S5). We also found that the abundance of species belongs to Actinobacteria (*Bifidobacterium longum*), Firmicutes (*Ruminococcus obeum* CAG 39 and *Holdemanella biformis*) and Proteobacteria (*Escherichia coli*) was significantly negatively correlated with ALT, AST and GGT levels, however, Firmicutes (*Coprococcus comes*) was significantly negatively correlated only with ALT and AST levels (P<0.05, Figure 3A, Dataset S5).

**Figure 3.**
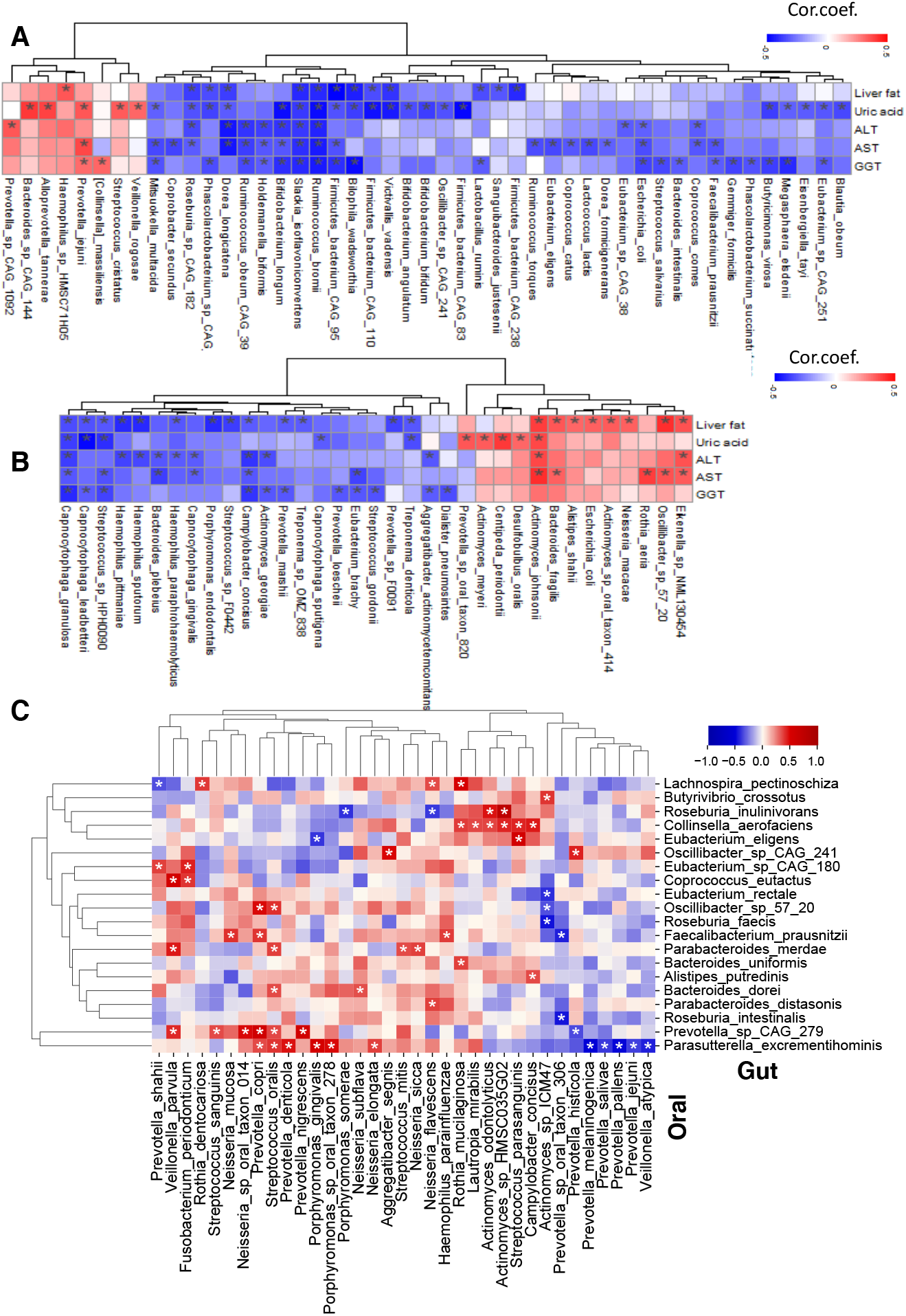
Heatmap shows the association between the five significantly different clinical variables’ plasma level with the abundance of species in the (A) gut microbiome and (B) oral microbiome. C) Heatmap shows the association between the abundance of species in the gut and oral microbiome. Asterisks indicate the statistical significance based on Spearman correlation with P<0.05. Cor.coef.: correlation coefficient.

The characterization of oral microbiome showed that the abundance of species belongs to Proteobacteria (*Campylobacter concisus*) and Bacteroides (*Capnocytophaga granulosa*) are negatively correlated with HS, ALT and AST levels. The abundance of species belongs to Proteobacteria (*Eikenella* sp NML130454) and Actinobacteria (*Actinomyces johnsonii*) are positively correlated with HS, ALT and AST levels (Figure 3B). Interestingly, we found that the reduced abundance in Haemophilus genus members (namely *H. paraphrohaemolyticus, H. pittmaniae* and *H. sputorum*) was negatively correlated with the HS and ALT levels. Comprehensively, AST and ALT levels were negatively correlated with the abundances of *Bacteroides plebeius* and *Capnocytophaga gingivalis;* HS and GGT were negatively correlated with the abundances of *Capnocytophaga leadbetteri, Streptococcus* sp HPH0090 and *Prevotella marshii*; HS and AST were positively correlated with the abundances of *Oscillibacte*r sp 57 20 and *Bacteroides fragilis*. Moreover, HS positively correlated with the abundances of *Alistipes shahii, Escherichia coli, Neisseria macacae, Actinomyces* sp oral axon 414 as well as negatively correlated with the abundances of *Porphyromonas endodontalis, Streptococcus sp F0442, Prevotella marshii, Treponema sp OMZ 838, Prevotella sp F0091* and *Treponema denticola* (P<0.05, Figure 3B, Dataset S5).

Several studies have reported that the gut microbiota plays a significant role in uric acid metabolism (Chiaro *et al*, 2017; Yu *et al*, 2018). We showed that the abundances of *Ruminococcus bromii, Slackia isoflavoniconvertens, Dorea longicatena, Firmicutes* bacterium CAG 95, *Firmicutes* bacterium CAG 110, *Bilophila wadsworthia, Victivallis vadensis, Roseburia* sp CAG 182 and *Phascolarctobacterium* sp CAG 266 in the gut microbiome and the abundances of *Capnocytophaga leadbetteri, Capnocytophaga granulosa, Streptococcus* sp HPH0090 and *Treponema denticola* in the oral microbiome were significantly negatively correlated with both uric acid levels and HS (P<0.05, Figure 3A & Figure 3B, Dataset S5). We also found that the abundances of *Bacteroides* sp CAG 144, *Alloprevotella tannerae, Prevotella jejuni, Streptococcus cristatus* and *Veillonella rogosae* in the gut microbiome and the abundances of *Centipeda periodontii, Prevotella* sp oral taxon 820, *Actinomyces meyeri* and *Desulfobulbus oralis* in the oral microbiome were significantly positively correlated with the uric acid levels (P<0.05, Figure 3A & Figure 3B, Dataset S5). We observed a negative correlation between uric acid levels and the abundances of species belongs to Actinobacteria (*Bifidobacterium angulatum, Bifidobacterium longum* and *Bifidobacterium bifidum*), Bacteroides (*Butyricimonas virosa*) and Firmicutes (*Mitsuokella multacida, Oscillibacter* sp CAG 241, *Firmicutes bacterium* CAG 83, *Megasphaera elsdenii, Blautia obeum, Eisenbergiella tayi* and *Eubacterium* sp CAG 251) in the gut microbiome (P<0.05, Figure 3A, Dataset S5), and the abundances of species belongs to Bacteroides (*Capnocytophaga sputigena*) in the oral microbiome (P<0.05, Figure 3B, Dataset S5)

### The link between the oral and gut microbiome

To study the transitions and interactions between the oral and gut microbiome, we performed correlation analysis between the abundance of species and observed significant correlations between them (P<0.05, Figure 3C, Dataset S6). We found that the abundance of the *Oscillibacter* sp CAG 241, which was significantly reduced in severe steatosis vs no steatosis and negatively associated with uric acid levels in the gut was significantly positively correlated with the abundance of *Prevotella histicola* and *Aggregatibacter segnis* in the oral microbiome. We also found that the abundance of the *Bacteroides uniformis*, significantly increased in moderate steatosis vs no steatosis, in the gut was significantly positively correlated with the abundance of *Rothia mucilaginosa* in the oral microbiome. Similarly, we found that the abundance of the *Roseburia inulinivorans* significantly increased in moderate steatosis vs no steatosis in the gut was significantly positively correlated with the abundance of *Actinomyces odontolyticus* and *Actinomyces sp* HMSC035G02 and significantly negatively correlated with the abundance of *Porphyromonas somerae* and *Neisseria flavescens* in the oral microbiome.

On the other hand, we found that the abundance of *Campylobacter concisus*, significantly negatively associated with HS in the oral microbiome was significantly positively correlated with the abundance of *Alistipes putredinis* and *Collinsella aerofaciens* in the gut microbiome (P<0.05, Figure 3C, Dataset S6). Moreover, we found that that the abundance of *Veillonella atypica*, significantly reduced in mild steatosis vs no steatosis in the oral microbiome was significantly negatively correlated with the abundance of *Parasutterella excrementihominis* in the gut microbiome. Of note, we found that abundances of *Parasutterella excrementihominis* in the gut microbiome were mostly affected by alterations in the abundance of different species in the oral microbiome (P<0.05, Figure 3C, Dataset S6).

We observed that the oral microbiota reflects the changes in the gut microbiota’s alterations in NAFLD patients. We also explored the associations between the abundance of oral and gut microbiome to study host and microbiome interactions in this context. Interest in butyrate-producing bacteria has increased with the earlier reports showing their essential role in the healthy human colon (La Rosa *et al*, 2019). Here, we found that the abundance of *Actinomyces* sp ICM47 in the oral microbiome was negatively correlated with butyrate-producing species, namely *Eubacterium rectale* and *Roseburia faecis* (P<0.05, Figure 3C, Dataset S6). Also, we found a negative correlation between the abundance of *Faecalibacterium prausnitzii* and *Roseburia intestinalis* in the gut microbiome with the abundance of *Prevotella* sp oral taxon 306 in the oral microbiome. Notably, there was a positive correlation between the abundance of *Faecalibacterium prausnitzii* in the gut microbiome with the abundance of *Haemophilus parainfluenzae, Neisseria mucosa* and *Prevotella copri* in the oral microbiome (P<0.05, Figure 3C, Dataset S6).

### Metabolomics alterations in the plasma of NAFLD patients

To study the interactions between the microbiome and host, we generated untargeted metabolomics data based on the plasma samples of the 56 subjects and measured the abundance of 1032 metabolites (Dataset S7). After excluding metabolites with missing values in >50% of samples, we analyzed the plasma level of 928 metabolites in the study (Dataset S8). Then, we identified the differentially expressed metabolites between groups and revealed the key metabolites associated with the underlying molecular mechanisms related to HS progression (Dataset S8).

We identified 43, 79 and 129 metabolites significantly differentially expressed in the mild, moderate, severe steatosis subject groups compared with no steatosis, respectively (Student’s t-test, P<0.05, Dataset S8). Of these metabolites, 17, 52 and 66 of them were associated with the lipid metabolism (Figure S1) whereas 26, 27 and 63 of them were related to other parts of metabolism (e.g. amino acids, NAD+ and antioxidant metabolism) (Figure 4A). Among these non-lipid metabolites, we found that 16 metabolites were significantly different only in mild steatosis vs no steatosis (Figure 4B), 14 metabolites significantly differed only in moderate steatosis vs no steatosis (Figure 4C), and 43 metabolites significantly differed only in the severe steatosis vs no steatosis (Figure 4D). We presented all 63 significantly different metabolites in the severe steatosis vs no steatosis in Figure 4D.

**Figure 4.**
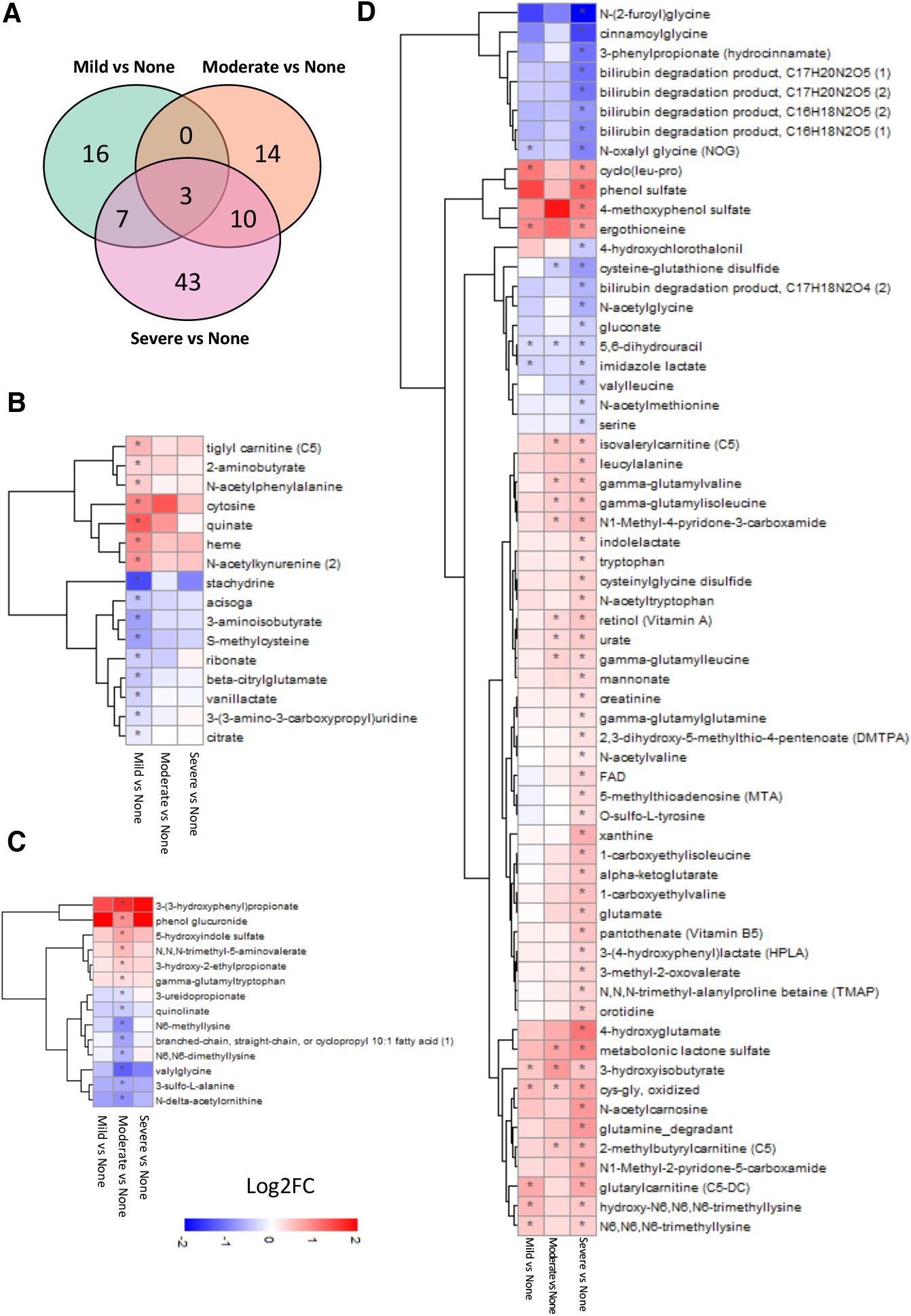
(A) Venn diagram shows significantly altered non-lipid metabolites in all groups compared to none steatosis. Heatmap shows Log2FC based alterations of metabolites that are exclusively different in the subjects with (B) mild steatosis and (C) moderate steatosis compared to the subjects with no steatosis. (D) All significantly altered non-lipid metabolites in the subjects with severe steatosis than the subjects with no steatosis are shown. Asterisks indicate statistical significance based on t-test. P<0.05. Log2FC: log2(fold change).

We observed that plasma level of 3-hydroxyisobutyrate (3-HIB), oxidized cys-gly were significantly higher and of 5,6-dihydrouracil was significantly lower in all the three steatosis groups vs no steatosis (Figure 4D). Previously, we measured plasma levels of 3-HIB, involved in branched-chain amino acids (BCAAs) metabolism in around 10,000 extensively phenotyped individuals, and identified 3-HIB as a marker of insulin resistance, mitochondrial dysfunction, and future risk of developing T2D (Mardinoglu *et al*, 2018b), which are closely linked to NAFLD. The oxidized form of l-cysteinylglycine is involved in glutathione metabolism, which has been reported to have a critical role in NAFLD’s progression and treatment (Mardinoglu *et al*, 2017; Mardinoglu *et al*., 2018a; Mardinoglu *et al*., 2018b; Mardinoglu *et al*., 2018c). The reduction of uracil produces 5,6-dihydrouracil, and it is involved in pyrimidine and beta-alanine metabolism. We found that the decreased plasma level of 5,6-dihydrouracil is associated with NAFLD.

We found the plasma level of heme was significantly higher only in mild steatosis vs no steatosis (Figure 4B). As the precursor of pro-or antioxidants of biliverdin and bilirubin, the alteration of heme synthesis may be associated with increased oxidative stress in NAFLD (Malaguarnera *et al*, 2005). Notably, we found that bilirubin’s degradation product was significantly lower in severe steatosis vs no steatosis (Figure 4D). N-acetyl kynurenine, which can promote inflammation (Rael *et al*, 2018), also increased only in mild steatosis vs no steatosis. The plasma level of quinolinate, a precursor for nicotinamide adenine dinucleotide (NAD^+^) synthesis, was significantly downregulated only in subjects with moderate steatosis vs no steatosis (Figure 4C). It has been reported that downregulation of quinolinate is associated with NAFLD in animals (Zeltser *et al*, 2020), and the altered NAD^+^ metabolism is associated with NAFLD in humans (Mardinoglu *et al*., 2017).

We found the plasma levels of serine, N-acetylglycine and glycine conjugated metabolites were significantly decreased in subjects with severe steatosis vs no steatosis (Figure 4D). Previously, we have found that NAFLD is associated with serine deficiency and reported that serine and glycine are key metabolites for glutathione synthesis (Mardinoglu *et al*, 2014), which is required for preventing the accumulation of intermediate products of fatty acid oxidation (Mardinoglu *et al*., 2017). We have proposed that serine supplementation may treat these patients (Zhang *et al*, 2020). We also found that the plasma level of cysteine-glutathione disulfide, a glutathione and cysteine conjugate product, was significantly lower in subjects with severe and moderate steatosis vs no steatosis. It has been reported that the concentration of cysteine-glutathione disulfide was significantly lower in subjects with steatosis and NASH (Kalhan *et al*, 2011). Moreover, we found all the bilirubin degradation products were downregulated only in severe steatosis vs no steatosis. Bilirubin can function as an antioxidant, reducing the HS accumulation (Hamoud *et al*, 2018; Kwak *et al*, 2012).

Moreover, we found that N,N,N-trimethyl-5-aminovalerate (TMAVA) plasma level was significantly increased in moderate steatosis vs no steatosis. The increase in the plasma level TMAVA in NAFLD patients has been shown recently, and it has been associated with the changes in the gut microbiome (Zhao *et al*, 2020). It has also been proposed as a predictor of microalbuminuria in patients with type 1 Diabetes (Haukka *et al*, 2018).

On the other hand, we found that plasma level of metabolites involved in tryptophan BCAA, lysine and uric acid metabolism was significantly increased in subject with severe steatosis (Figure 4D). We observed that the plasma level of uric acid and xanthine involved purine metabolism was significantly increased in subjects with severe steatosis vs no steatosis. We also found that the plasma level of N,N,N-trimethyl-alanylproline betaine (TMAP) associated with urea cycle was significantly increased in subjects with severe steatosis vs no steatosis. These results agree with the previous studies, where plasma uric acid level is significantly associated with HS in NAFLD patients (Li *et al*, 2009). Although the metabolites mentioned above were significantly different in one or two HS groups, most of these metabolites had a tendency to decrease or increase by following the direction of differences in other HS groups.

### Associations between the plasma metabolome and patient phenotype

We assessed the associations between the plasma level of significantly different five clinical parameters including liver fat, uric acid, ALT, AST and GGT with the plasma level of metabolites (Figure 5A, Dataset S9). We found that all these clinical variables were positively correlated with the plasma level of TMAP and BCAA and its conjugates, including leucine, isoleucine, gamma-glutamyl leucine, gamma-glutamyl isoleucine, gamma-glutamyl valine, 3-methyl-2-oxovalerate and N-acetylcarnosine. Like 3-HIB, other BCAAs products have been associated with NAFLD progression, insulin resistance, mitochondrial dysfunction and incidence of T2D (Honda *et al*, 2017; Yoon, 2016). The level of ALT, AST and GGT was positively correlated with the plasma level of kynurenine, can activate inflammatory response and has been associated with NAFLD (Rael *et al*., 2018). Moreover, uric acid’s plasma level was significantly positively correlated with all these parameters.

**Figure 5.**
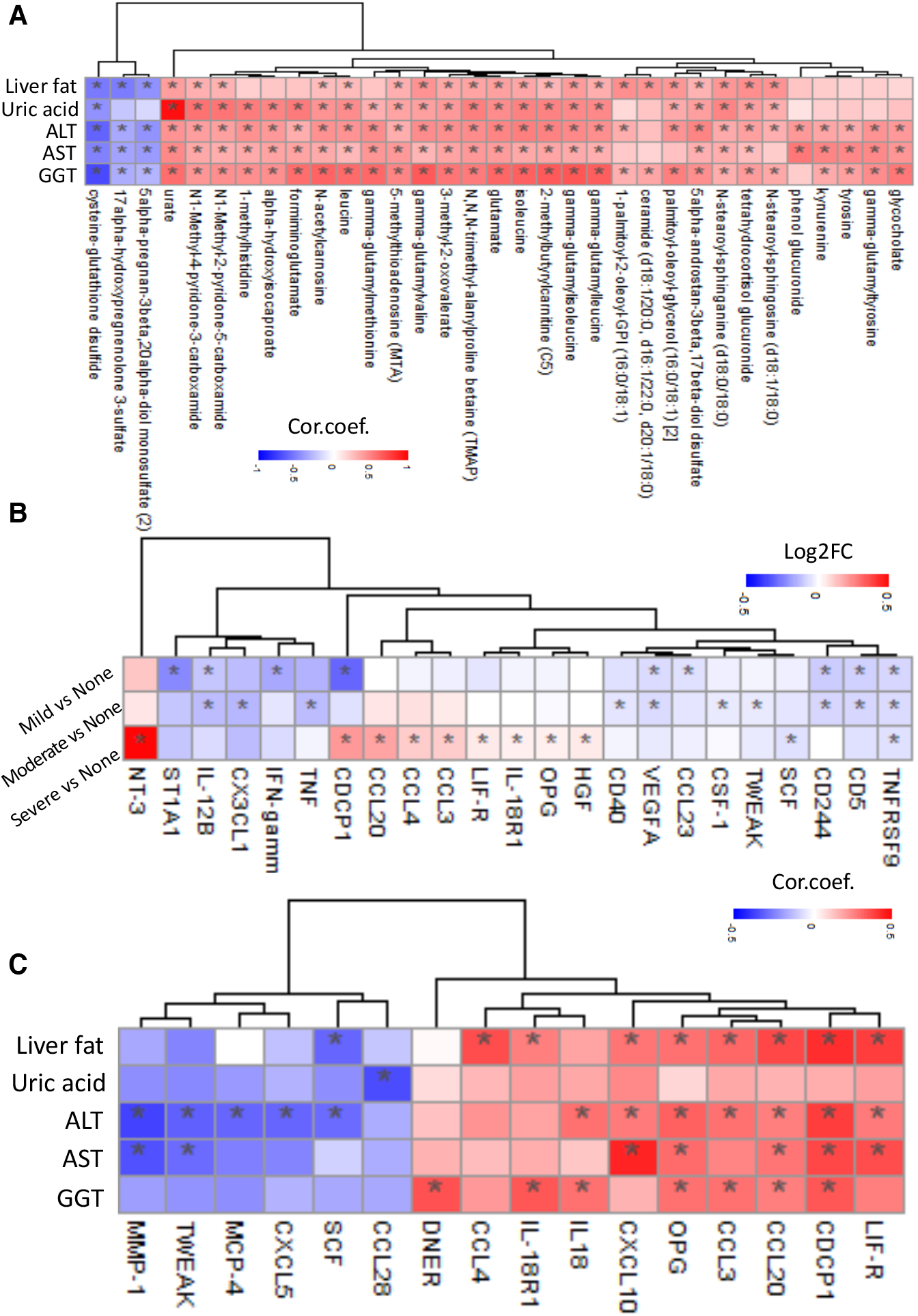
(A) Heatmap shows the association between the five significantly different clinical variables’ plasma level with the top 10 most significantly correlated metabolites’ plasma level. Asterisks indicate the statistical significance based on Spearman correlation with P<0.05. (B) Heatmap shows the Log2FC based alterations of all the significantly altered inflammation-related proteins in the subjects with mild, moderate and severe steatosis compared to the subjects with no steatosis. Asterisks indicate the statistical significance based on t-test with P<0.05. (C) Heatmap shows the association between the plasma level of the five significantly different clinical variables with the inflammation-related proteins. Asterisks indicate the statistical significance based on Spearman correlation. P<0.05. Log2FC: log2(fold change). Cor.coef.: correlation coefficient

In contrast, cysteine-glutathione disulfide’s plasma level was significantly negatively correlated with these 5 clinical parameters. The plasma level of 17alpha-hydroxypregnenolone 3-sulfate and 5alpha-pregnan-3beta,20alpha-diol monosulfate (2) was significantly negatively associated with HS, ALT, AST and GGT levels (Figure 5A). The differences in these metabolites’ plasma level can be used to detect HS and explore the effect of treatment in NAFLD patients.

### The influence of the microbiome on the plasma metabolome

In the gut microbiome, the abundances of the species belong to Bacteroidales (*Alistipes finegoldii, Alistipes putredinis, Bacteroides dorei, Bacteroides stercoris, Bacteroides uniformis, Bacteroides vulgatus, Parabacteroides distasonis, Parabacteroides merdae*) are significantly positively correlated with numerous lipid structures and negatively correlated with the secondary bile acid metabolites, specific peptides and amino acids (namely gamma-glutamyl-alpha-lysine, S-adenosylhomocysteine, gamma-glutamyl threonine and threonine) (Figure S2, Dataset S13). Similarly, the abundances of the species belong to Clostridiales (*Butyrivibrio crossotus, Coprococcus eutactus, Eubacterium eligens, Eubacterium* sp CAG 180, *Faecalibacterium prausnitzii, Lachnospiraceae pectinoschiza, Oscillibacter* sp 57 20, *Oscillibacter* sp CAG 241, *Roseburia faecis, Roseburia intestinalis*) are negatively correlated with primary bile acid metabolites as well as numerous peptides and amino acids (Figure S2, Dataset S13). Interestingly, *Provetolla copri* displayed an opposite behaviour than other Bacteroidales, and it is negatively correlated with those lipid structures but positively correlated with the amino acids (Figure S2, Dataset S13). Similarly, *Eubacterium rectale*, a member of Clostridiales, positively correlated with primary bile acid metabolism and negatively correlated with long-chain polyunsaturated fatty acids (Figure S2, Dataset S13). Hence, we observed that the abundance *Bacteroides uniformis*, significantly increased in subjects with moderate steatosis vs no steatosis and *Oscillibacter* sp CAG 241 that was significantly reduced in subjects with severe steatosis vs no steatosis is significantly correlated with the plasma metabolites involved in bile acid and amino acid metabolism.

In the oral microbiome, the correlation between the abundances of the individual species and significantly altered plasma metabolites in different HS groups including phenol sulfate (tyrosine metabolism) was positively correlated with the abundances of *Prevotella shahii, Prevotella denticola, Porphyromonas gingivalis, Porphyromonas somerae, Porphyromonas* sp oral taxon 278 and *Neisseria subflava* but negatively correlated with the abundances of the *Actinomyces odontolyticus, Actinomyces* sp HMSC035G02 and *Rothia dentocariosa*. Moreover, the plasma level of phenol glucuronide (tyrosine metabolism) was positively correlated with the abundances of *Prevotella* sp oral taxon 306, *Porphyromonas gingivalis* and *Prevotella intermedia* but negatively correlated with the abundances of *Rothia dentocariosa* and *Streptococcus sanguinis* (Figure S3, Dataset S13). We also observed that the abundance of *Campylobacter concisus*, significantly negatively correlated with HS and *Veillonella atypica*, significantly reduced in subjects with mild steatosis vs no steatosis was significantly correlated with the glutamate and phenol sulfate, respectively. These species’ abundances were also associated with the plasma level of metabolites involved in amino acid metabolism, carnitine metabolism, and lipid metabolism.

The plasma level isovalerylcarnitine, associated with BCAA metabolism, was positively correlated with the abundances of *Alloprevotella tannerae, Prevotella nigrescens* and *Prevotella oulorum* but negatively correlated with the abundances of *Prevotella histicola* and *Streptococcus infantis*. The plasma level of tiglyl carnitine, associated with BCAA metabolism, was positively correlated with the abundances of *Alloprevotella tannerae* and *Prevotella shahii* but negatively correlated with the abundances of *Streptococcus parasanguinis, Megasphaera micronuciformis, Veillonella atypica, Veillonella dispar* and *Neisseria sicca*.

The plasma level of N6,N6,N6-trimethyllysine (lysine metabolism) was positively correlated with the abundances of *Porphyromonas gingivalis* and *Porphyromonas somerae* but negatively correlated with the abundances of *Veillonella atypica, Prevotella histicola, Prevotella salivae* and *Neisseria* sp oral taxon 014. The plasma level of 2,3-dihydroxy-5-methylthio-4-pentenoate (methionine, cysteine, and taurine metabolism) was positively correlated with the abundances of *Prevotella nigrescens* but negatively correlated with the abundances of *Actinomyces odontolyticus, Actinomyces* sp HMSC035G02, *Prevotella intermedia* and *Campylobacter concisus*. The plasma level of glutamate (glutamate metabolism) was positively correlated with the abundances of *Prevotella copri* and *Neisseria mucosa* but negatively correlated with the abundances of *Prevotella intermedia, Fusobacterium periodonticum* and *Campylobacter concisus*; urate (purine metabolism) was positively correlated with the abundances of *Prevotella oulorum, Prevotella* sp oral taxon 306, *Tannerella* sp oral taxon HOT 286 but negatively correlated with the abundances of P*orphyromonas endodontalis* and *Prevotella intermedia*. All correlations between individual species in the oral microbiome and plasma metabolites presented in Figure S3 and Dataset S13.

### Inflammatory proteomics alterations in the plasma of NAFLD patients

Plasma levels of 94 inflammatory protein markers were measured by the proteome profiling platform Proximity Extension Assay (PEA). After quality control and exclusion of proteins with missing values in more than 50% of samples, 72 proteins were analysed (Dataset S10). Proteins whose expression levels significantly differed between groups were presented in Dataset S11.

Except the plasma level of CDCP1 which is significantly lower in mild steatosis and significantly higher in severe steatosis vs no steatosis, majority of the proteins followed the same directional changes in all steatosis groups (Figure 5B). It has been reported that CDCP1 knockout mice have increased lipid accumulation in the liver (Wright, 2017) which may explain the downregulation of CDCP1 in mild steatosis compared with no steatosis in our study. Besides, CDCP1 also acts as a profibrotic mediator which may play a central role in subjects with severe steatosis and fibrosis (Noskovičová *et al*, 2018).

We found that the plasma level of TNFRSF9 was significantly lower in all three steatosis groups vs no steatosis (Figure 5B). It has been reported that stimulation of TNFRSF9 with agonistic antibody reduces adiposity, body weight and HS and increases energy expenditure in diet-induced obese mice and genetically obese/diabetic mice (Seijkens *et al*, 2014). The plasma level of ST1A1, IFN-gamma and CCL23 was lower only in mild steatosis vs no steatosis (Figure 5B). The differences in the mRNA expression of ST1A1 is associated with high-fat diet-induced obesity. The plasma levels of CX3CL1, TNF, CD40, CSF-1 and TWEAK were lower in moderate steatosis vs no steatosis (Figure 5B). The downregulation of CX3CL1/CX3CR1 pathway has been suggested as a mechanism underlying β cell dysfunction in type 2 diabetes (Lee *et al*, 2013). It has been reported that TNF level contributes HS in diet-induced obesity (De Taeye *et al*, 2007), CD40 deficiency in mice exacerbates obesity-induced HS and insulin resistance (Guo *et al*, 2013), mice lacking CSF-1 have reduced adiposity (Ferrante, 2007) and decreased serum level of TWEAK concentration is associated with the NAFLD(Willberg *et al*, 2007).

The plasma level of NT-3, CCL20, CCL4, CCL3, LIF-R, OPG and HGF were higher, and SCF was lower only in the severe steatosis vs no steatosis (Figure 5B). It has been shown that the protein level of CCL20 was increased in NAFLD (Chu *et al*, 2018; Hanson *et al*, 2019). An increased mRNA expression of LIF-R has been demonstrated in high-fat diet-induced NAFLD mice (Toye *et al*, 2007). Moreover, increased circulating levels of HGF have been reported in NASH (Balaban *et al*, 2006).

Then we assessed the associations of the significantly differed five clinical variables with the plasma levels of the inflammation-related proteins (Figure 5C, Dataset S12). We identified two main clusters. These variables were negatively and positively correlated with at least one of the inflammation-related proteins in the first and second clusters, respectively.

### The influence of the microbiome on the plasma proteome

In the gut microbiome, we found that the abundances of *Coprococcus eutactus, Dialister* sp CAG 357, *Oscillibacter* sp 57 20 and *Eubacterium* sp CAG 180 were most positively associated with the inflammatory protein levels (Figure S4, Dataset S14). On the other hand, the abundances of *Roseburia intestinalis, Eubacterium eligens, Parabacteroides distasonis, Roseburia faecis, Butyrivibrio crossotus* and *Prevotella copri* were negatively correlated with the plasma levels of inflammation-related proteins (Figure S4, Dataset S14). Of note, the IL10 plasma level was positively correlated with the abundances of *Collinsella aerofaciens* and *Alistipes finegoldii* but negatively correlated with the abundances of *Roseburia intestinalis*, a primary degrader of dietary fiber (La Rosa *et al*., 2019) (Figure S4, Dataset S14).

In the group with the moderate steatosis; we found that CSF1 was positively correlated with the abundance of *Parasutterella excrementihominis*; TWEAK was positively correlated with the abundance of *Oscillibacter* sp CAG 241; CCL23 was negatively correlated with the abundances of *Roseburia intestinalis, Eubacterium eligens, Barnesiella intestinihominis* and *Roseburia faecis* (Figure S4, Dataset S14). Additionally, associations of significant proteins in the group with severe steatosis with gut microbiome revealed a negative correlation between HGF plasma level and the abundances of *Butyrivibrio crossotus* and *Roseburia intestinalis*; a negative correlation between NT-3 plasma level and the abundances of *Prevotella* sp CAG 279; a positive correlation between OPG plasma level and the abundances of *Dialister* sp CAG 357 and *Coprococcus eutactus*; a positive correlation between CCL3 plasma level and the abundances of Oscillibacter sp 57 20, *Dialister* sp CAG 357 and *Coprococcus eutactus*; a positive correlation of CCL4 plasma level and the abundances of *Roseburia inulinivorans* and *Coprococcus eutactus*, but a negative correlation with the abundances of *Roseburia intestinalis*; a positive correlation of CCL20 plasma level and the abundances of *Coprococcus eutactus* (Figure S4, Dataset S14). In brief, our findings suggested that the abundances of *Coprococcus eutactus* are positively correlated with the levels of the inflammatory proteins that are significantly altered in subjects with severe steatosis.

In the oral microbiome, we found species within *Neisseria* genus (*N. mucosa, Neisseria sp oral taxon 014, N. elongate, N. subflava, N. sicca*), *Rothia* genus (*R. aeria, R. dentocariosa, R. mucilaginosa*) and *Veillonella* genus (*V. parvula, V. atypica*) were positively associated with the numerous inflammatory proteins (Figure S5, Dataset S14). However, there was a negative correlation between the abundance of species belonging to the *Porphyromonas* genus (namely *P. somerae, P. endodontalis, P. gingivalis*) and the *Prevotella* genus (namely *P. pallens, P. oulorum, P. shahii, P. intermedia*) with the inflammation-related proteins (Figure S5, Dataset S14). Interestingly, the abundances of the *Neisseria flavescens, Haemophilus parainfluenzae* and *Campylobacter concisus* were also negatively correlated with inflammation-related proteins (Figure S5, Dataset S14). Besides, FGF-21 plasma level was negatively correlated with the abundances of *Streptococcus mitis* and *Tannerella* sp oral taxon HOT 286; CDCP1 plasma level was positively correlated with the abundances of *Neisseria mucosa* and negatively correlated with the abundances of *Tannerella* sp oral taxon HOT 286; IL-6 plasma level was negatively correlated with the abundances of *Porphyromonas endodontalis* and CSF-1 plasma level was negatively correlated with the abundances of *Alloprevotella tannerae* (Figure S5, Dataset S14). Other significantly correlated species with plasma inflammation-related proteins are presented in Figure S5, Dataset S14.

### Prediction of HS based on multi-omics data

We analysed phenomics, metabolomics, proteomics and oral/gut metagenomics data and identified features differentiating between the groups of subjects with varying HS degrees. We performed analyses using single/multi-omics data using the Random Forest algorithm and discovered the key features associated with HS. First, we used all individual data points from each omics data (Figure 6A, Figure S6 & S7, Dataset S15). We found that the gut metagenomics data was the top-performing dataset in the prediction of the steatosis degree, with > 70% accuracy (AUC: 0.90) (Figure 6A). In contrast, we found that the inflammatory proteomics data was the worst-performing data, with only 35.3% accuracy (AUC: 0.72) (Figure 6A). Next, we tested the top 5 or 10 features from each omics dataset and tried their different combinations (Figure 6B, Dataset S15 & Figure S8A-H). We observed that the varieties of top features in clinical (5 variables), metabolomics (10 variables), and proteomics (5 variables) yielded 64.7% accuracy (AUC: 0.92) Figure S8A-F), whereas adding gut and oral metagenomics (10 top features each) showed the highest accuracy of 94.1% (AUC: 0.951) in the prediction of the steatosis degree (Figure 6B & Figure S8G-I). In the validation dataset, the model yielded 82.7% accuracy (AUC: 0.843) by adding these top features in clinical (5 variables), metabolomics (10 variables), and proteomics (5 variables) and gut and oral metagenomics (10 top features each) (Figure 6C & Figure S8J). We had the highest predictivity of HS (AUC: 1.0) by adding top 5 features in clinical, metabolomics, proteomics, gut metagenomics and oral metagenomics data in the finding dataset (Figure 6D). We also validated the predictivity of the final model with 22 subjects and found that the model yielded a higher predictivity AUC: 0.886) (Figure 6E) with the same key features compared to other combinations (Figure S8J).

**Figure 6.**
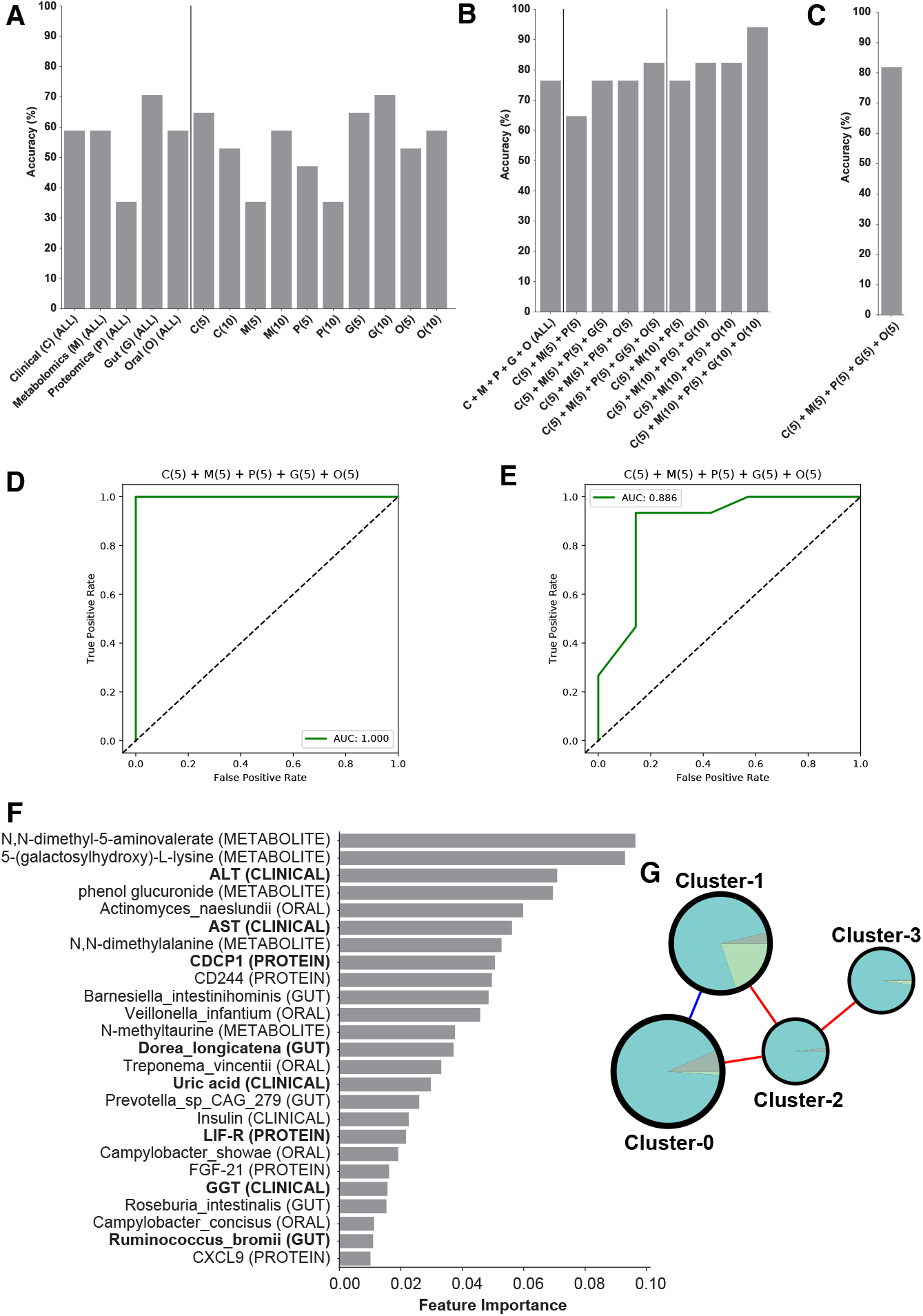
Accuracy score of random forest classification algorithm performed to predict the class using single omics (A) and multi-omics combination of top features from each omics type in finding (B) and validation datasets (C). Numbers in the brackets represent the number of top features taken from the single omics data. (D) AUC-ROC curve for prediction of HS based on the data from 56 subjects. (E) AUC-ROC curve for validation of the final model in prediction of HS based on the data from 22 subjects. (F) Top 25 features from the model with the highest accuracy, C(5) + M(5) + P(5) + G(5) + O(5). Analytes that are altered significantly between the groups are marked in bold. (F) Composition of each cluster in the multi-omics network.

The top 5 features in clinical (ALT, AST, uric acid, insulin and GGT), metabolomics (N,N-dimethyl-5-aminovalerate, 5-(galactosylhydroxy)-L-lysine, phenol glucuronide, N,N-dimethylalanine and N-methyltaurine), proteomics (CDCP1, CD244, LIF-R, FGF-21 and CXCL9), gut metagenomics (*Barnesiella intestinihominis, Dorea_longicatena, Prevotella* sp CAG_279, *Roseburia_intestinalis* and *Ruminococcus_bromii*) and oral metagenomics (*Actinomyces naeslundii, Veillonella infantium, Treponema vincentii, Campylobacter showae* and *Campylobacter concisus*) used in prediction of HS can be considered as candidate biomarkers for NAFLD (Figure 6F, Dataset S15). In our study, we also found that the abundance of the gut microbiome species including *Barnesiella intestinihominis, Dorea longicatena* and *Ruminococcus bromii* and oral microbiome species including *Campylobacter concisus* and *Veillonella infantium* was significantly associated with HS and reported that their abundance was significantly correlated with the plasma level of metabolites and inflammatory proteomics levels.

Some of these markers have been reported and validated in other studies. In our previous study (Lovric *et al*, 2018), we found that the level of ALT, AST, Insulin, CDCP1, and FGF-21 were significantly positively correlated with the various ectopic fat depots, including HS. Moreover, the abundance of gut microbiome species, including *Dorea Logicatena* and R*uminococcus bromii*, have been reported as possible markers to predict HS (Brahe *et al*, 2015; Lee *et al*, 2020). The abundance of oral microbiome species, including *porphyromonas endodontalis* and *Campylobacter concisus*, was also significantly negatively correlated with the HS based on previous studies.

### Integrative analysis of multi-omics data using biological networks

We generated an integrative multi-omics network for showing the relationships between different analytes within and between other omics datasets. The network was built using Spearman correlation analysis (Dataset S16), between all analytes from the aforementioned omics data, filtered by edges with FDR < 0.05, resulting in a relatively sparse network with 1032 nodes and 17536 edges (3.3% network density). The complete network is presented in iNetModels (http://inetmodels.com), an interactive multi-omics networks database and visualization. We performed a centrality analysis on the network by calculating each node’s degree (Dataset S16). We found that the top 20 most connected nodes were related to lipid metabolism, e.g. ceramides, sphingomyelins, diacylglycerol and phospholipid-related sub-pathways. Similarly, we found lipid-related clinical variables as top nodes in clinical variables (Total cholesterol, GGT, LDL, and TG-level). In contrast, top inflammatory proteomics nodes were STAMBP, TNFSF14, SIRT2, CXCL5, CXCL1 and CD40, associated with cytokine-cytokine receptor interaction, and NF-kappa B, TNF, and IL-17 which are associated with several signalling pathways.

Subsequently, we performed a clustering analysis using the Leiden community analysis algorithm. We then combined the smaller clusters (11 clusters with 1-54 analytes) to the next biggest cluster (cluster-3) to balance the cluster size, resulting in 4 clusters to use in further analysis (Figure 6G, Dataset S16). We found that all clusters had positive correlations with each other, except the connection between cluster-0 and cluster-1 (Figure 6G), based on their shared edges. Looking at the central analytes in each cluster (Dataset S14), as expected due to the unbalanced number of analytes in the clusters, metabolites dominate each cluster’s top analytes. In cluster-0, metabolites related to amino acid metabolisms were on top, specifically isoleucine and it’s derivative and gamma-glutamyl amino acids, together with a clinical variable, GGT, whereas cluster-1 was dominated by phospholipid, carbohydrate, and taurine metabolism, followed by the central proteins. Top 20 most connected metabolites and top clinical variables mentioned above, were concentrated in cluster-2, which contained most lipid-related metabolites and clinical variables, making this cluster as candidate central variables in HS (Dataset S16). Finally, we found that cluster-3 contained mostly metabolites related to fatty acid metabolism. Interestingly, the clustering analysis showed sub-networks with analytes, which has similar functionality; this shows the power of biological networks in establishing the functional relationships between analytes based on the multi-omics analysis.

We took the features from our multi-omics random forest model and filtered only the significantly altered analytes, resulting in 9 analytes fulfilling those requirements (ALT, AST, GGT, Uric acid, CDCP1, LIF-R, *Dorea longicatena* (gut), *Ruminococcus bromii* (gut), *Porphyromonas endodontalis* (oral)). Next, we retrieved the subnetwork with the first neighbour of those features as well as HS and overlaid comparative analysis results (filtering P < 0.05) (Figure 7A). We found that those key features related with HS, and their first neighbours were dominated by lipid metabolites, gamma-glutamyl amino-acids, BCAA metabolites, fatty acid metabolism (carnitine derivatives), and glutathione-related metabolites that are significantly associated with the HS.

**Figure 7.**
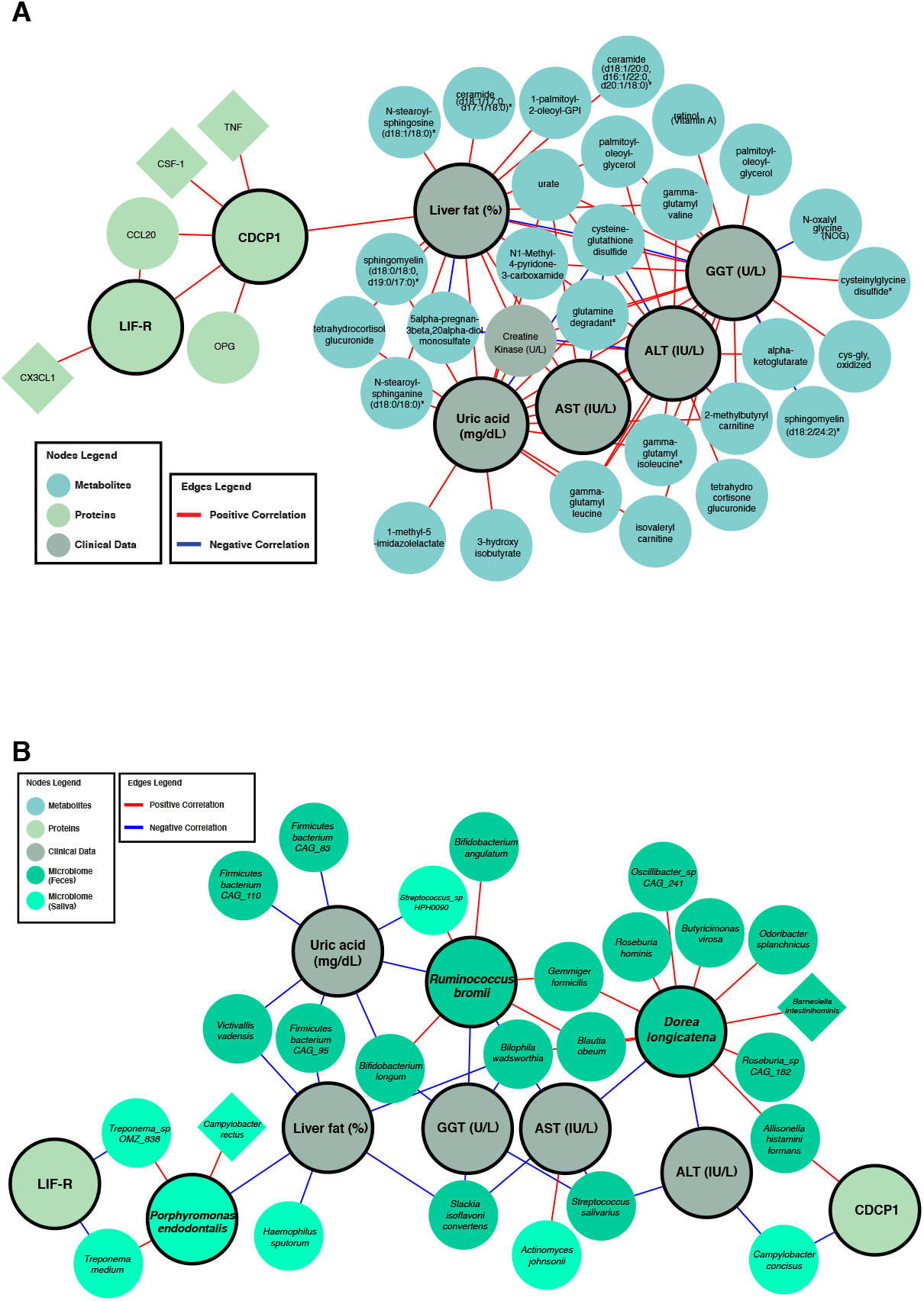
Significantly different first neighbours of significantly different analytes in the best performing random forest model in the (A) multi-omics and (B) metagenomics-centric correlation networks.

Metagenomics data were not included in the multi-omics network built previously due to the data’s sparsity that caused dissociation of the network hence reducing the network analysis’s power. We decided to construct a metagenomics centric network, by including the key features from random forest analyses and all microbial species with > 1% abundance in at least five samples (Figure 7B). We have integrated the complete networks into iNetModels platform. Similar to the previous approach, we overlapped the comparative analysis results (P<0.05) and retrieved the sub-network. HS was significantly negatively correlated with the abundance of *Victivallis vadensis, Firmicutes bacterium* CAG 95, *Slackia isoflavoniconvertens*, and *Bilophila wadsworthia* in the gut, and *Porphyromonas endodontalis* and *Haemophilus sputorum* in the oral microbiome. Interestingly, the abundance of *Slackia isoflavoniconvertens*, and *Bilophila wadsworthia* in the gut also negatively correlated with HS, AST and GGT levels. Another key species in the gut microbiome is *Streptococcus salivarius*, whose abundance was negatively correlated with ALT, AST and GGT levels. On the other hand, the abundance of *Dorea longicatena* was negatively correlated with both ALT and AST levels and positively correlated with numerous members of Bacteroidales and Clostridiales. We also found that the abundance of *Campylobacter concisus* is associated with levels of both ALT and CDCP1, a transmembrane receptor associated with aggressive epithelial cancers. Additionally, we observed a positive correlation between AST levels and the abundance of *Actinomyces johnsonii* in the oral microbiome. We also observed that abundance of *Ruminococcus bromii* was significantly positively associated with the level of uric acid and GGT. Uric acid levels are also negatively correlated with the abundance of specific species in Firmicutes phylum and *Bifidobacterium longum* and *Victivallis vadensis*. Based on these results, the integrative network analysis with multi-omics data strengthened the results from single omics analyses and added additional power to identify key features associated with HS. Moreover, it allowed us to reveal functional relationships within and between different omics data.

## Discussion

The microbial imbalance has been identified as a key player in NAFLD’s pathogenesis due to the functional crosstalk between liver and complex microbial composition (Altay *et al*., 2019). Microbiota can improve or aggravate liver diseases through several mechanisms, including enhanced liver lipid metabolism, elevated alcohol production, altered energy metabolism, impaired intestinal permeability and disrupted bile secretion (Guohong *et al*, 2019; Safari & Gérard, 2019). Previous studies demonstrated that NAFLD patients had higher TNF-alpha and IL-6 in the mucosal layer of the intestinal wall (Jiang *et al*, 2015) and reported larger quantities of pathogenic bacteria in the gut (Aron-Wisnewsky *et al*, 2020).

Our study used shotgun sequencing (enabling enhanced taxonomic resolution) of saliva and faeces samples to analyse the composition of oral and gut microbiota. We showed that these diverse communities are associated with different steatosis levels in a well-characterized overweight and obese NAFLD cohort. In the gut microbiome, we identified significant alterations in certain species following the existing literature; emphasizing that the abundances of *Dorea longicatena* were reduced in patients with steatosis, and the abundances of *Slackia isoflavoniconvertens, Roseburia hominis* and *Ruminococcus bromii* were reduced in severe steatosis. Some of these species have been proposed to be essential for healthy microbiota homeostasis previously. The abundances of *Dorea longicatena* has been found to be reduced in NAFLD-cirrhosis (Loomba *et al*., 2017), negatively correlated with the markers of insulin resistance in postmenopausal women with obesity (Brahe *et al*., 2015) and exhibited higher levels in remission of Crohn’s disease (Mondot *et al*, 2016). The abundances of *Slackia isoflavoniconvertens*, is an equol producer bacteria by conversion of the soy isoflavone, has been endorsed as having many favourable effects on the host metabolism (Mayo *et al*, 2019); *Ruminococcus bromii* is another beneficial species for human health, and its abundance was inversely correlated with the fibrosis severity and primary bile acid levels in non-obese NAFLD subjects. An interventional animal study suggested a potential role in synthesising secondary bile acids (Lee *et al*., 2020). Moreover, the abundance of *Barnesiella intestinihominis* was found to be significantly overrepresented in the stool with a potency to induce NAFLD based on 16S rDNA profiling of mice(Le Roy *et al*, 2013).

Generation of the metabolomics and proteomics data allowed for studying the molecular pathways and identifying key features associated with the NAFLD progression. We observed that the metabolites involved in the glutathione metabolism, BCAA metabolism, and pyrimidine metabolism, which are the key pathways in NAFLD, were already altered from HS’s early stage. In the patients with moderate steatosis, we identified TMAVA, a biomarker used to predict gut microbiome change, confirming that TMAVA may be an essential feature of NAFLD (Zhu *et al*, 2020). We found that plasma level of TMAVA was significantly positively correlated with the abundance of *Bacteroides stercoris, Bacteroides uniformis, Parabacteroides distasonis* and negatively correlated with the abundance of *Prevotella copri* in the gut microbiome. The TMAVA plasma level is also significantly negatively correlated with the abundance of the *Veillonella dispar* and *Veillonella atypica*, one of the key species associated with HS, in the oral microbiome. Notably, we identified N,N-dimethyl-5-aminovalerate (di-methylated forms TMAVA) as one the most critical feature in the prediction of NAFLD.

Moreover, we observed that serine and glycine related metabolites were altered in the severe stage of steatosis, further highlighting their crucial roles in NAFLD (Mardinoglu *et al*., 2014; Mardinoglu *et al*., 2017). Besides, we observed that heme, the precursor of antioxidant of bilirubin, and bilirubin degradation products were altered in mild and severe steatosis compared with no steatosis, respectively. This suggests that the redox balance may be changed at the early stage of HS. Based on the comparison of proteomics data of different degrees of steatosis vs no steatosis, we observed a decreasing tendency of most inflammation-related proteins in mild and moderate vs no steatosis but an increasing trend in the severe steatosis vs no steatosis.

More commonly, the liver immune tolerance mechanism, processing immunosuppressive functions by regulating cytokines or chemokines’ expression, limits the magnitude of intrahepatic immune responses and allows the liver to recover (Crispe, 2014). However, immune tolerance is broken by the further accumulation of fat, which induces severe steatosis. As a result, enhanced antigen presentation to lymphocytes associated with the increased expression of inflammation-related proteins leads to the development of both cellular and humoral immune responses (Sutti & Albano, 2020). Besides, we observed that the abundance of the species, including *Barnesiella intestinihominis, Oscillibacter* sp CAG 241 and *Roseburia inulinivorans* associated with HS was significantly correlated with the inflammatory proteomics plasma levels. We also observed that the abundance of HS associated species including *Campylobacter concisus* (negatively correlated with CXCL9), *Porphyromonas endodontalis* (negatively correlated with LIF-R) and *Veillonella atypica* (positively correlated with CD244) in the oral microbiome was significantly correlated with the plasma level of the inflammatory proteomics plasma levels.

In conclusion, we performed a multi-omics analysis of subjects with varying degrees of HS and integrated these data using systems approaches to identify HS’s key features. We revealed the alterations in the microbial compositions start at early stages of the clinical spectrum and cause metabolic disturbances underlying HS. We also studied the effect of these alterations on the host metabolism by performing plasma metabolomics, and inflammatory proteomics analysis. Hence, we revealed the underlying molecular mechanisms involved in the progression of HS. We envisage that our results can be used to discover prognostic and predictive clinical markers and develop efficient therapeutic strategies.

## Materials and Methods

### Participants

Overweight or obese patients 18–70 years of age were enrolled in the trial if they were diagnosed with NAFLD and met all the inclusion criteria: body mass index >27 kg/m^2^, triglycerides ≤354 mg/dl, low-density lipoprotein cholesterol ≤175 mg/dl, and increased HS (>5.5%). Patients were excluded if they carried the PNPLA3 I148M (homozygous for I148M), had ALT or AST levels >3-fold higher than the upper limit of normal, or had taken oral antidiabetics, including metformin, within three months. The main characteristics of the study participants are presented in Dataset S1.

MRI-PDFF determined HS, and plasma samples for proteomics and metabolomics analyses were collected (Dataset S1). Patients for this characterization study were recruited at the Koç University Hospital, Istanbul, Turkey (Dataset S1). The study was conducted following Good Clinical Practice guidelines and the principles of the Declaration of Helsinki. An independent external data monitoring committee oversaw the safety of the participants and the risk-benefit analysis. Written informed consent was obtained from all participants before trial-related procedures were initiated. The Koç University ethics committee approved the study (Decision No: 2018.351.IRB1.043, Decision Date: 15 May 2019).

### Metagenomics Data Analysis

Fresh stool and saliva specimens were collected and preserved using DNA/RNA Shield Fecal Collection tubes (Zymo Research, Irvine, CA) and DNA/RNA Shield Collection Tube (Zymo Research, Irvine, CA), respectively. DNA extractions from the faecal samples were done using QIAamp PowerFecal Pro DNA Kit (Qiagen, Hilden, Germany) and the saliva samples using QIAamp DNA Microbiome Kit (Qiagen, Hilden, Germany). All protocol procedures were performed according to the manufacturer’s instructions. Quantification of extracted DNA was determined fluorometrically on the Qubit® 3.0 Fluorometer (Thermo Fisher Scientific, United States) using the QubitTM dsDNA HS Assay Kit. DNA purity was determined via 260/280 and 260/230 ratios measured on the NanoDrop 1000 (Thermo Fisher Scientific, United States). The SMARTer Thruplex DNA-Seq (Takara Bio) was used for library preparation (Option:350 bp; Category: low input). Samples were sequenced on NovaSeq6000(NovaSeq Control Software 1.7.0/RTA v3.4.4) with a 151nt (Read1)-10nt(Index1)-10nt(Index2)-151nt(Read2) setup using ‘NovaSeqXp’ workflow in ‘S4’ mode flow cell. The Bcl to FastQ conversion was performed using bcl2fastq_v2.20.0.422 from the CASAVA software suite. The quality scale used is Sanger /phred33/Illumina 1.8+.

Raw paired-end metagenomics data were analysed using Metaphlan3 (Beghini *et al*, 2020) to extract each sample’s taxonomic profiles. The abundant data were then analysed using the Wilcoxon rank-sum test to identify the species different between subjects with no steatosis compared to the other groups. Spearman correlation analysis was used to analyse the associations between selected analytes and the taxonomic abundance data. The correlation between oral and gut metagenomics data (by filtering the species with abundance > 1% in at least 5 data points). The *SciPy* package was used. All analyses were done using Python 3.

### Untargeted Metabolomics Analysis

Plasma samples were collected for nontargeted metabolite profiling by Metabolon (Durham, NC). The samples were prepared with an automated system (MicroLab STAR, Hamilton Company, Reno, NV). For quality control purposes, a recovery standard was added before the first step of the extraction. To remove protein and dissociated small molecules bound to protein or trapped in the precipitated protein matrix, and to recover chemically diverse metabolites, proteins were precipitated with methanol under vigorous shaking for 2 min (Glen Mills GenoGrinder 2000) and centrifuged. The resulting extract was divided into four fractions: one each for analysis by ultraperformance liquid chromatography-tandem mass spectroscopy (UPLC-MS/MS) with positive ion-mode electrospray ionization, UPLC-MS/MS with negative ion-mode electrospray ionization, and gas chromatography-mass spectrometry; one fraction was reserved as a backup.

### Inflammatory Protein Markers

In the plasma samples, inflammatory protein markers were determined with the Olink Inflammation panel (Olink Bioscience, Uppsala, Sweden). Briefly, each sample was incubated with 92 DNA-labeled antibody pairs (proximity probes). When an antibody pair binds to its corresponding antigens, the corresponding DNA tails form an amplicon by proximity extension, which can be quantified by high-throughput, real-time PCR. Probe solution (3 μl) was mixed with 1 μl of sample and incubated overnight at 4°C. Then 96 μl of extension solution containing extension enzyme and PCR reagents for the pre-amplification step was added. The extension products were mixed with detection reagents and primers and loaded on the chip for qPCR analysis with the BioMark HD System (Fluidigm Corporation, USA). To minimize inter and intra-run variation, the data were normalized to both an internal control and an interplate control. Normalized data were expressed in arbitrary units (Normalized Protein eXpression, NPX) on a log2 scale and linearized with the formula 2^NPX^. A high NPX indicates a high protein concentration. The limit of detection, determined for each of the 92 assays, was defined as three standard deviations above the negative control (background).

### Statistical Analysis

Values are expressed as the mean ± standard deviation (SD) (continuous variables) or as n (%). For all analyses, metabolites and proteins that were missing in > 50% of patients were removed. Each subject group was compared with subjects with no steatosis with *t-test* from the *Scipy* package. Missing values were dropped before the analysis. Principal component analysis (PCA) was performed using *scikit-learn* package. Finally, Spearman correlation analysis was used to analyze the association between selected analytes and other datasets (metabolomics and proteins). The *SciPy* package was used. All analyses were done using Python 3.7.

### Random Forest Analysis

A random forest classification algorithm was used to find each dataset’s key features and each network cluster. The analyses were performed using the *RandomForestClassifier* function from the *scikit-learn* package. Several trees were calculated before the analysis by selecting the highest accuracy with the lowest number of trees (up to 100 trees). Sample bootstrapping and out-of-bag sample options were enabled. AUC-ROC was generated using the same module, by combining the classes (no and mild steatosis, and moderate and severe steatosis)

### Generation of Multi-Omics Network

Multi-omics network was generated based on the Spearman correlations and the significant associations (FDR < 0.05). The analyses were performed with the *SciPy* package in Python 3.7. Centrality analysis and Leiden Clustering (community analysis) on the network were performed using *iGraph* Python and *leidenalg* module. Missing values were omitted in a pairwise manner (using *nan_policy = ‘omit’* option). Networks were visualized using Cytoscape 3.8.2. All networks presented on this manuscript can be access openly in iNetModels (http://inetmodels.com).

## Code Availability

All code used for the analyses is available in https://github.com/sysmedicine/nafldBaseline.

## Data Availability

All data associated with this study are available in the main text or the supplementary materials.

## SUPPLEMENTAL DATA

Supplemental Information includes 8 supplementary figures and 16 supplementary datasets.

## ACKNOWLEDGMENTS

This work was financially supported Knut and Alice Wallenberg Foundation. The authors would like to thank the Plasma Profiling Facility team at SciLifeLab in Stockholm for generating the Olink proteomics data, Metabolon Inc. (Durham, USA) for the generation of metabolomics data and NGI, Scilifelab for the generation of metagenomics data.

The authors gratefully acknowledge the use of the services and facilities of the Koç University Research Center for Translational Medicine (KUTTAM), equally funded by the Republic of Turkey Ministry of Development Research Infrastructure Support Program. Findings, opinions or points of view expressed in this article do not necessarily represent the official position or policies of the Ministry of Development.

The computations were performed on resources provided by SNIC through Uppsala Multidisciplinary Center for Advanced Computational Science (UPPMAX) under Project sens2019031.

## Contributors

Study Design, M.U., J.B., A.M.; Patient Recruitment, M.Z., M.A., B.S., M.G.G.; Investigation, H.Y., O.A., M.A., C.F., W.K., X.L., J.M.S., H.T., C.Z., S.S., J.N.; Data Analyses, O.A., M.A., C.F., W.K., X.L., J.M.S., C.Z., S.S., J.N., A.M.; Interpretation, M.Z., M.U., J.B., A.M.; Drafting the manuscript, M.Z, O.A., A.M.; Revising the manuscript, all authors.

## SUPPLEMENTARY FIGURE LEGENDS

**Figure S1** (A) Venn diagram shows significantly altered lipid metabolites in all comparisons. Heatmap shows Log2FC based alterations of metabolites that are exclusively different in the subjects with (B) mild steatosis and (C) moderate steatosis compared to the subjects with no steatosis. (D) All significantly altered lipid metabolites in the subjects with severe steatosis compared to the subjects with no steatosis. Asterisks indicate statistical significance based on t-test. P<0.05. Log2FC: log2(fold change).

**Figure S2** Heatmap is showing the Spearman correlation score (paired) between metabolites and gut microbiota species (abundance > 1%). Asterisks denotes significant correlations (P < 0.05). Only metabolites that were significantly correlated with 5 or more species are shown in the heatmap

**Figure S3** Heatmap is showing the Spearman correlation score (paired) between metabolites and oral microbiota species (abundance > 1%). Asterisks denotes significant correlations (P < 0.05). Only metabolites that were significantly correlated with 5 or more species are shown in the heatmap

**Figure S4** Heatmap is showing the Spearman correlation score (paired) between proteins and gut microbiota species (abundance > 1%). Asterisks denotes significant correlations (P < 0.05).

**Figure S5** Heatmap is showing the Spearman correlation score (paired) between proteins and oral microbiota species (abundance > 1%). Asterisks denotes significant correlations (P < 0.05).

**Figure S6** Top 20 features from (A) clinical, (B) metabolomics, and (C) proteomics data identified by random forest classification. Analytes altered significantly (P < 0.05) between severe vs no steatosis comparison are marked in bold.

**Figure S7** Top 20 features from (A) Gut and (B) Oral microbiome data identified by random forest classification. Analytes altered significantly (P < 0.05) between severe vs no steatosis comparison are marked in bold.

**Figure S8 (**A) – (I) AUC-ROC curves for HS prediction based on single/multi-omics data based on the data from 56 subjects. (J) AUC-ROC curve for validation of the final model predicts HS based on the data from 22 subjects.

## SUPPLEMENTARY DATASETS

Dataset S1 Clinical and physical variables (A-C) are presented for the 56 subjects, recruited in the finding cohort. D) The mean values for these variables and the differences in the clinical and physical variables between subjects with mild, moderate, and severe hepatic steatosis are compared to those with no hepatic steatosis.

Dataset S2 Clinical and physical variables (A-C) and the mean values (D) of the variables are presented for the 22 subjects, recruited in the validation cohort. Multi-omics data, including oral (E) and gut (F) metagenomics, metabolomics (G) and proteomics (H) data generated for the 22 subjects.

**Dataset S3 Metagenomics Raw Data for each of 56 subject recruited in the study.**

Dataset S4 Differences in the abundance of the species in the gut and oral microbiome between the subjects with mild, moderate and severe hepatic steatosis compared to those with no hepatic steatosis.

Dataset S5 Associations between the abundance of the species in the gut and oral microbiome and the level of significantly altered clinical variables are presented.

Dataset S6 Association between the abundances of species in the gut and oral microbiome is presented.

**Dataset S7 Untargeted metabolomics data for each of 56 subject recruited in the study.**

Dataset S8 Differences in plasma level of metabolites between the subjects with mild, moderate and severe hepatic steatosis compared to the subjects with no hepatic steatosis. Only metabolites detected in >50% of samples were analysed.

Dataset S9 Associations between the plasma level all metabolites and the level of significantly altered clinical and physical variables are presented.

**Dataset S10 The Olink multiplex inflammation panel used to detect the dynamic range of 72 proteins in the subjects’ plasma samples.**

Dataset S11 Differences in the plasma level of inflammation-related proteins between the subjects with mild, moderate and severe hepatic steatosis compared to the subjects with no hepatic steatosis. Only proteins detected in >50% of samples were analysed.

Dataset S12 Associations between the plasma level of all inflammation-related proteins and the level of significantly altered clinical and physical variables are presented.

Dataset S13 Associations between the plasma level all metabolites and the abundance of the species in the gut and oral microbiome are presented.

Dataset S14 Associations between the plasma level all proteins and the abundance of the gut and oral microbiome species are presented.

Dataset S15 Highly ranked metabolites, proteins, species and clinical features based on Random Forest analysis.

Dataset S16 Multi-Omics Network Data, including edges and nodes information, are presented. The network is shown in the iNetModels (http://inetmodels.com).

