## Supplementary figures and images for "Multi-omics analysis reveals the impact of microbiota on host metabolism in hepatic steatosis"

### Supplementary Figure 1.pdf

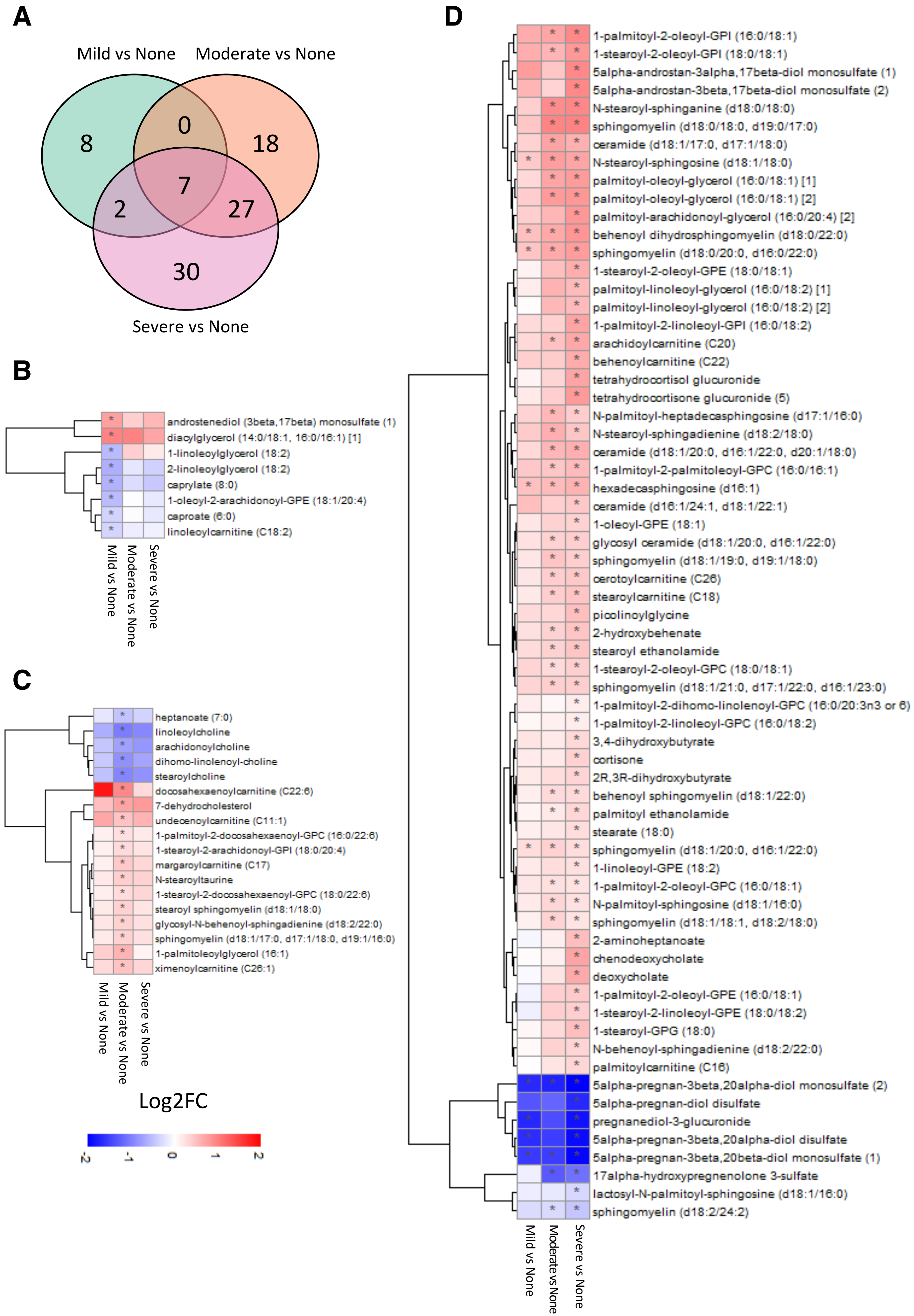

### Supplementary Figure 2.pdf

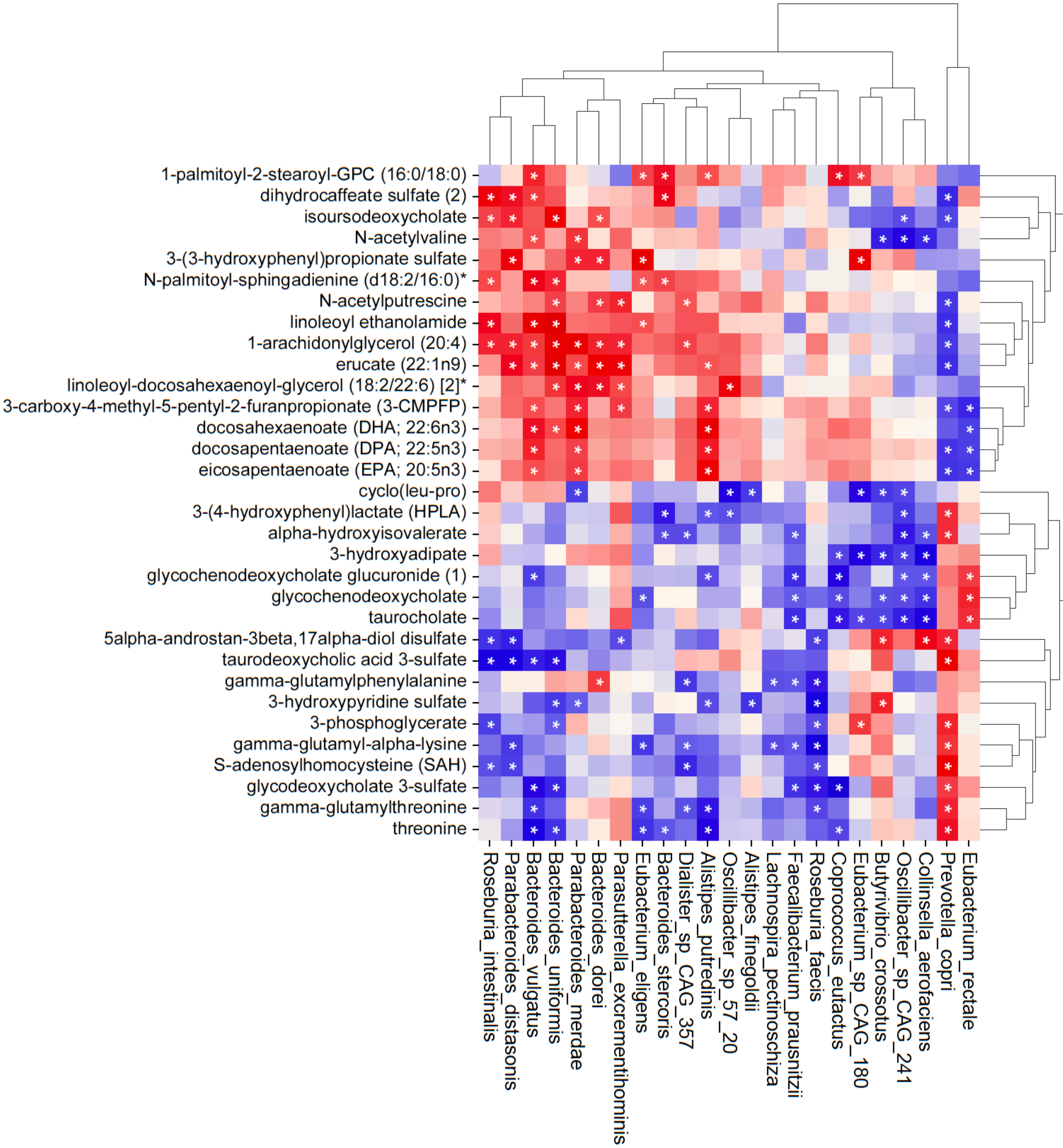

### Supplementary Figure 3.pdf

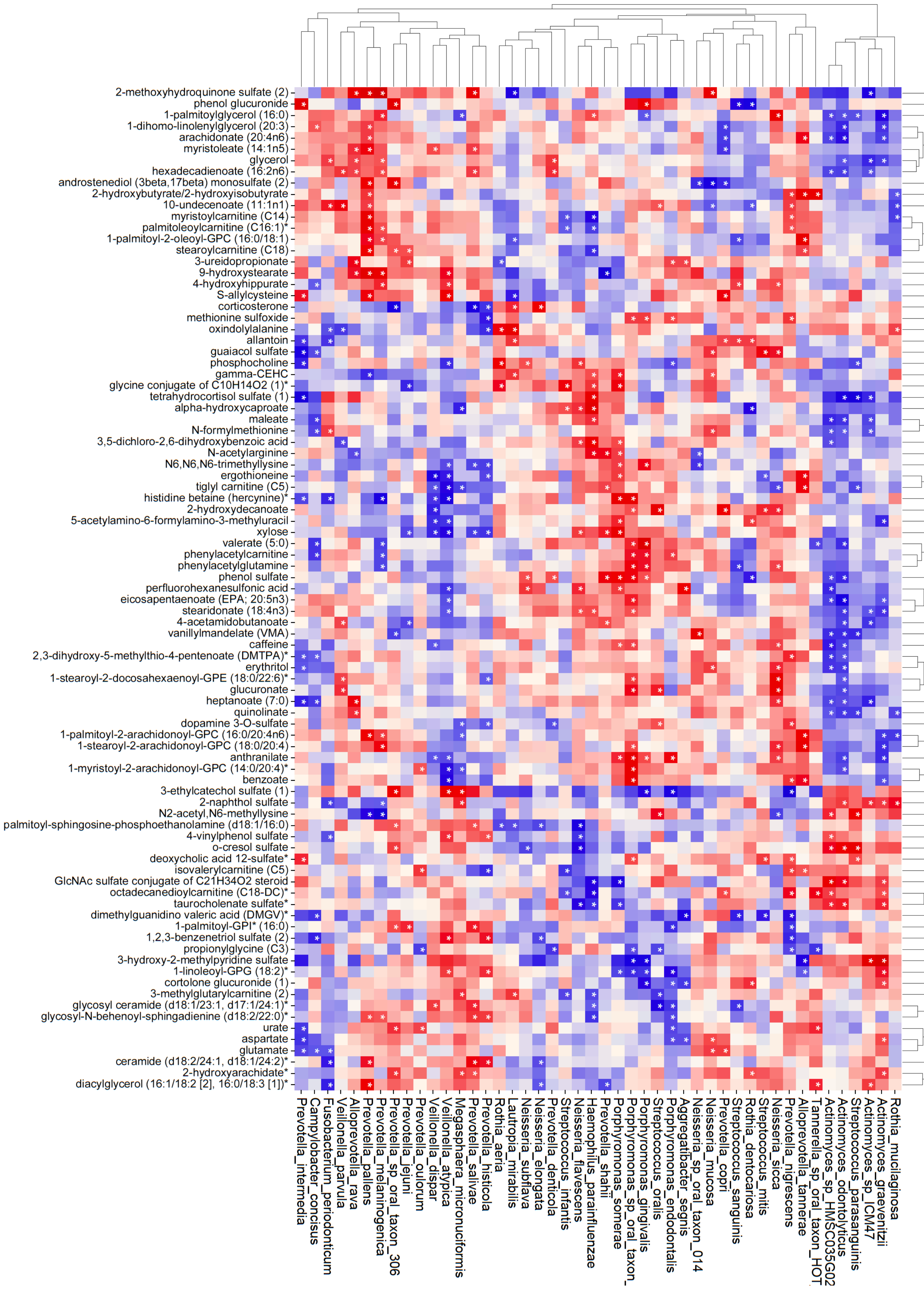

### Supplementary Figure 4.pdf

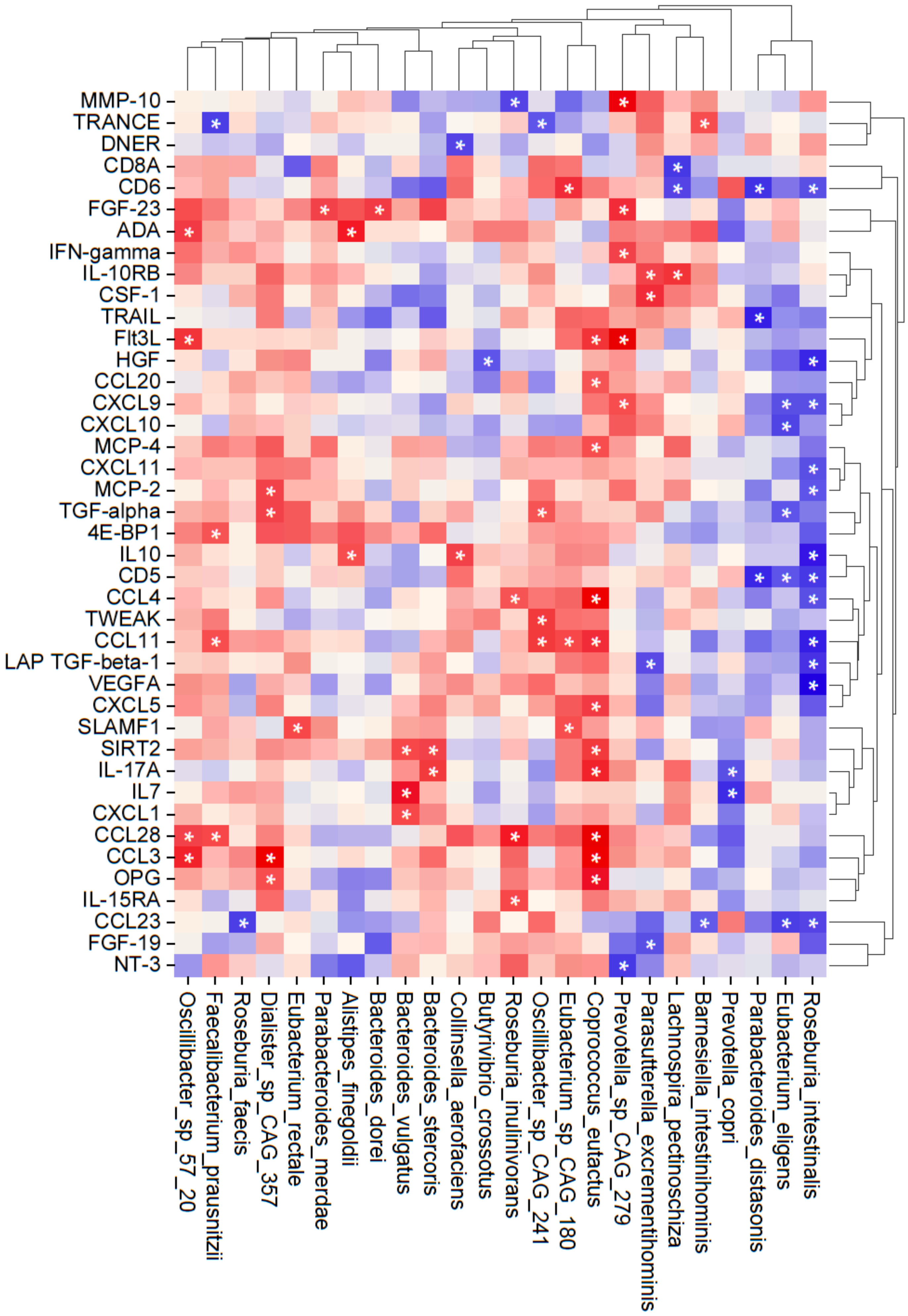

### Supplementary Figure 5.pdf

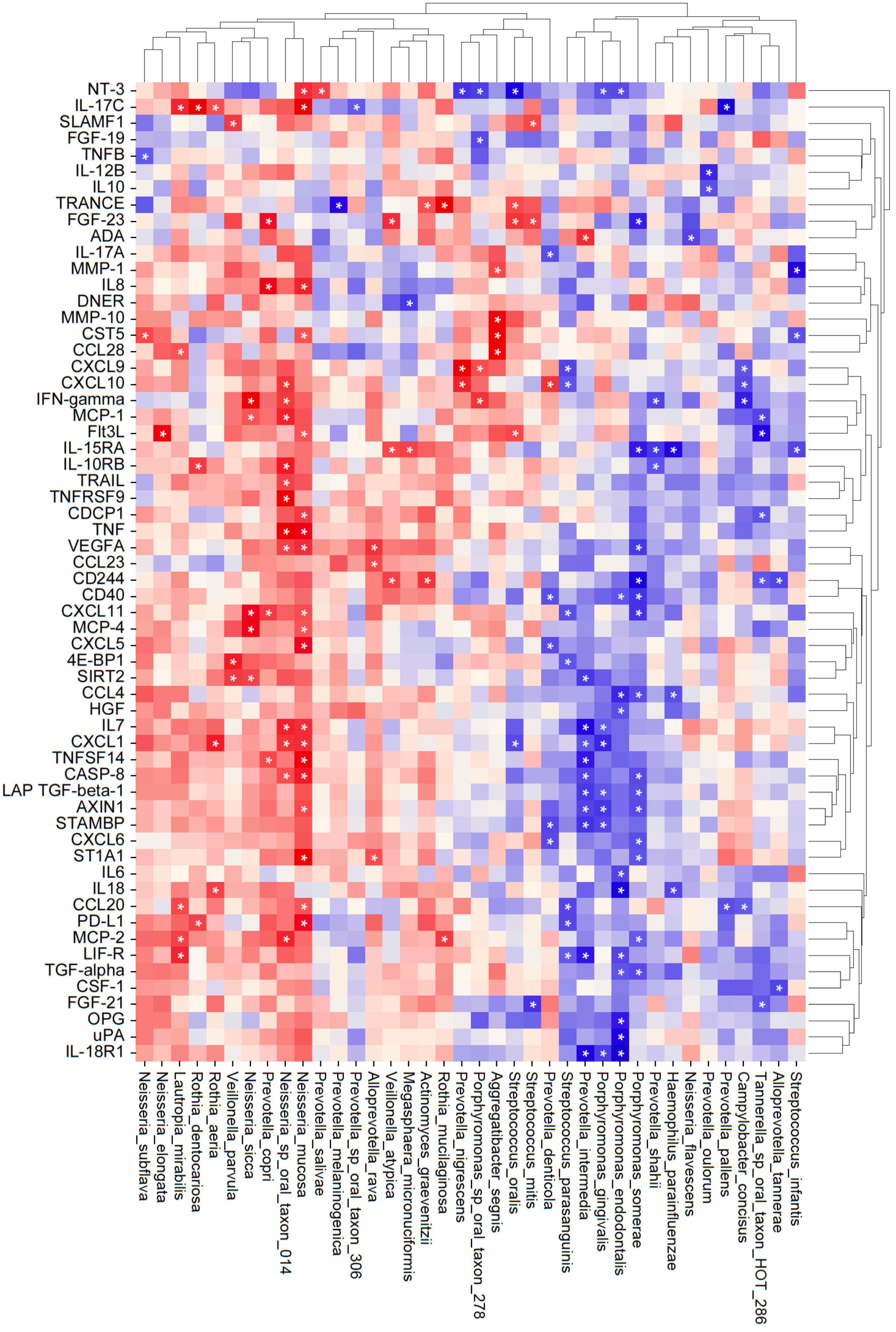

### Supplementary Figure 6.pdf

#### A) Clinical Data

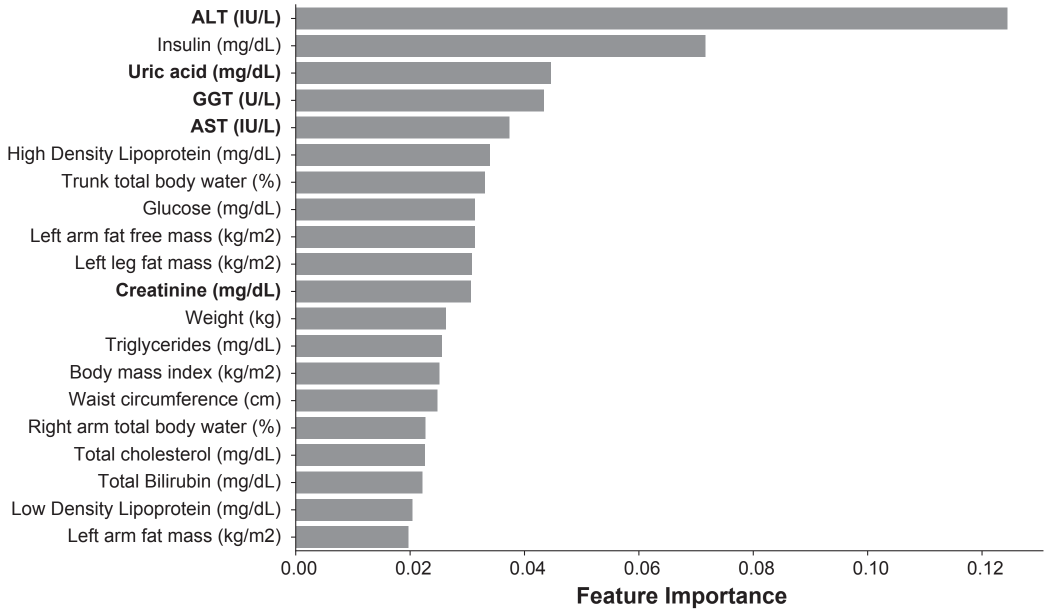


#### B) Metabolomics

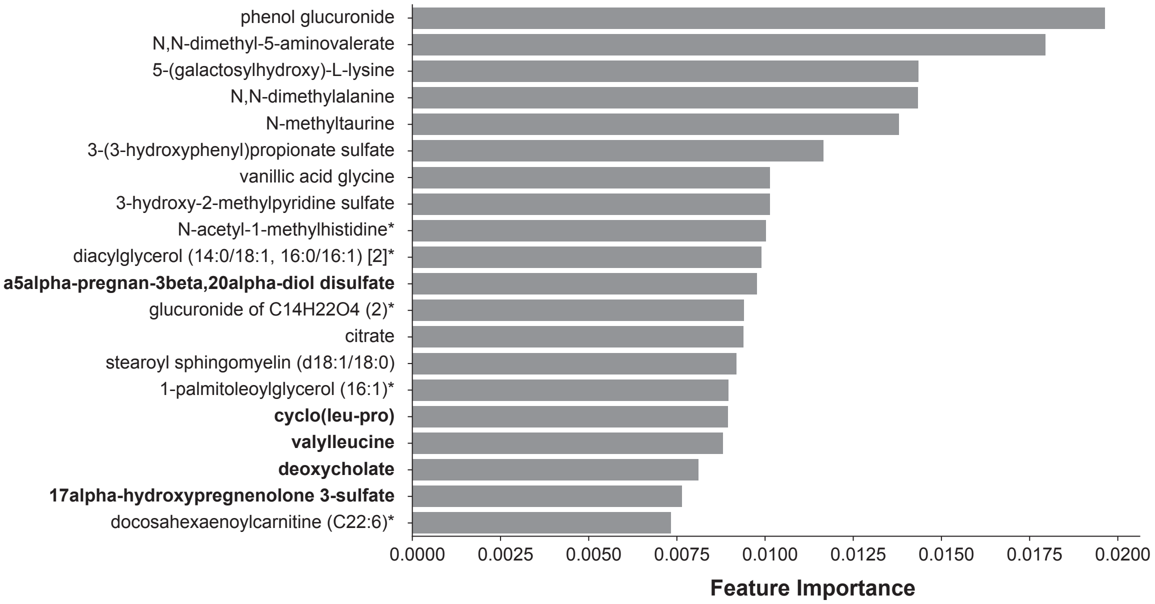


#### C) Proteomics

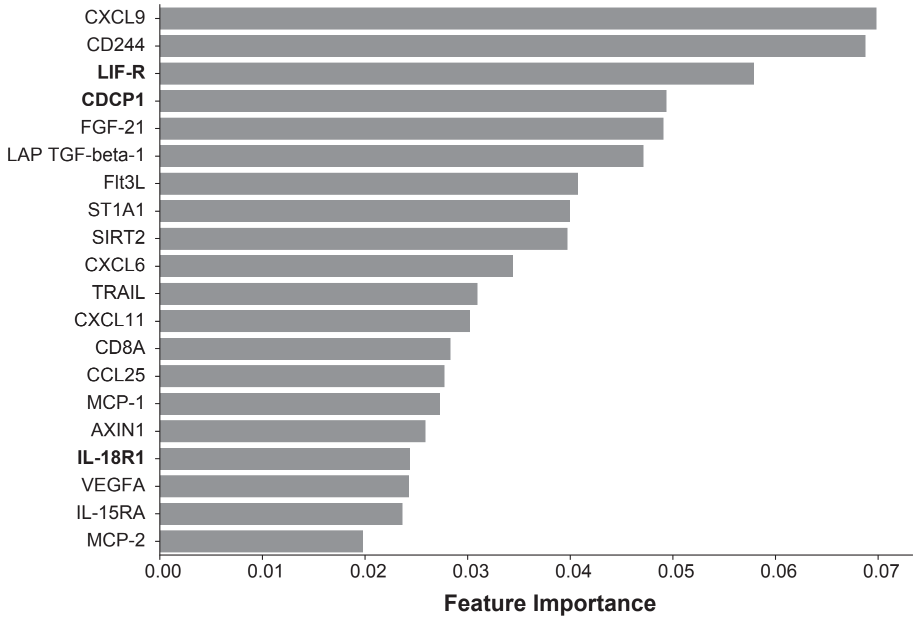

### Supplementary Figure 7.pdf

#### A) Gut Microbiome

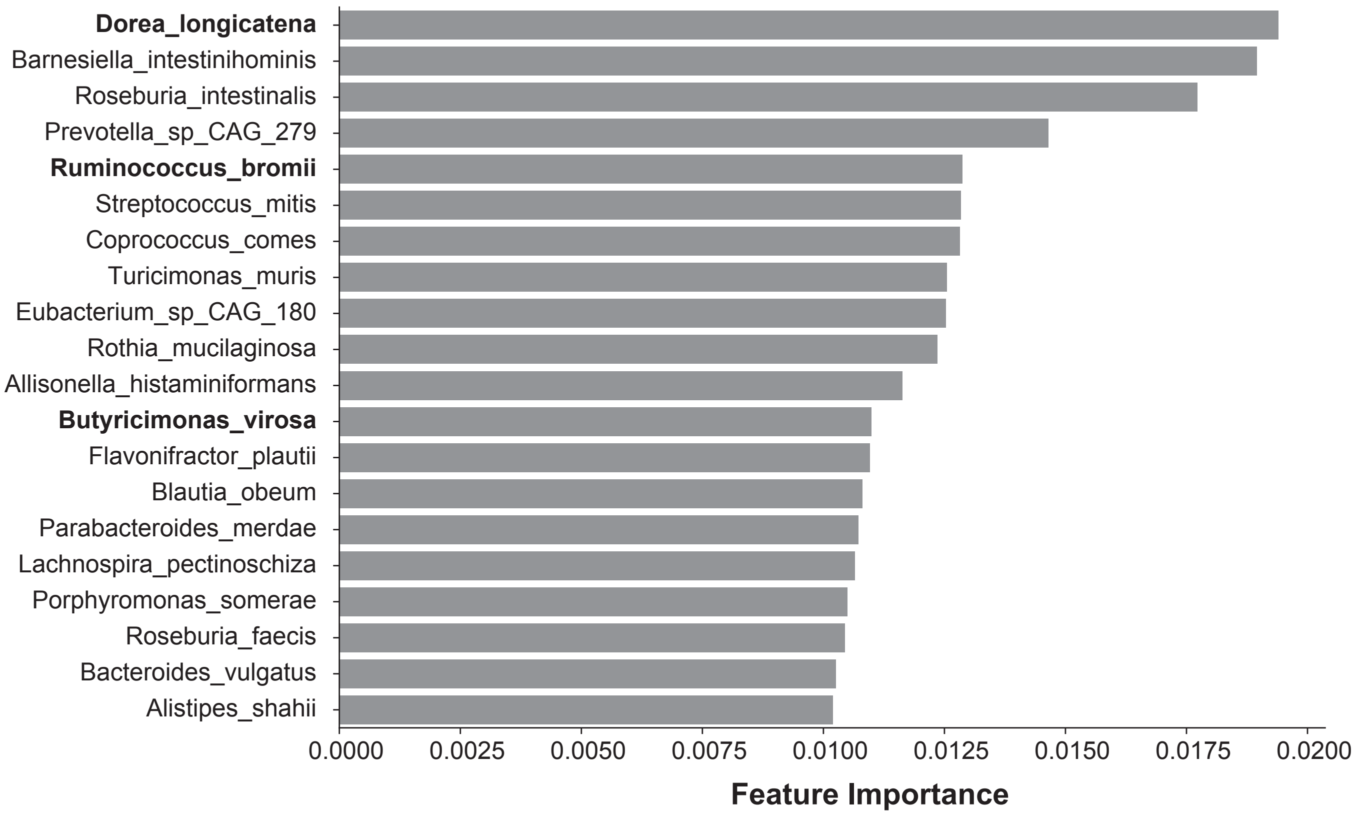


#### B) Oral Microbiome

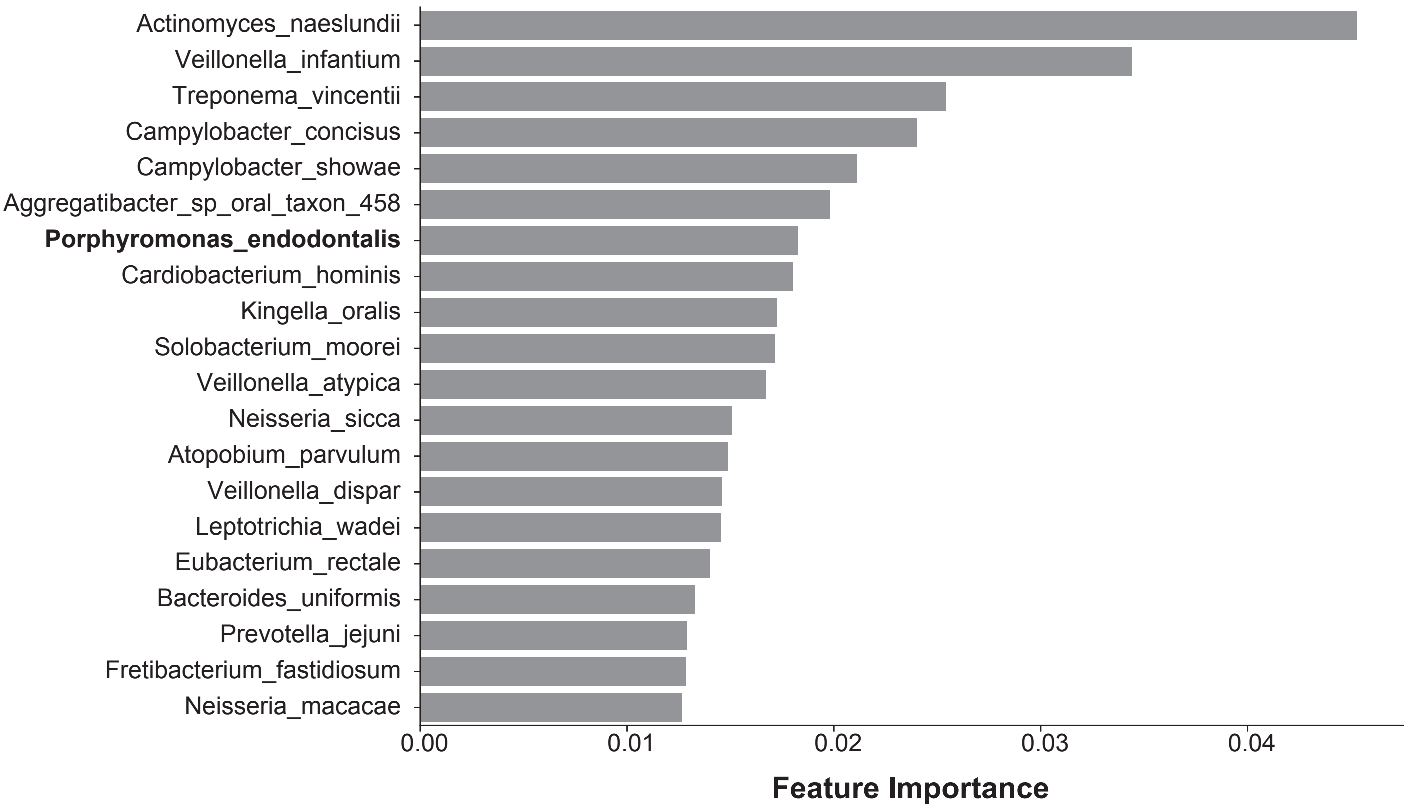

### Supplementary Figure 8.pdf

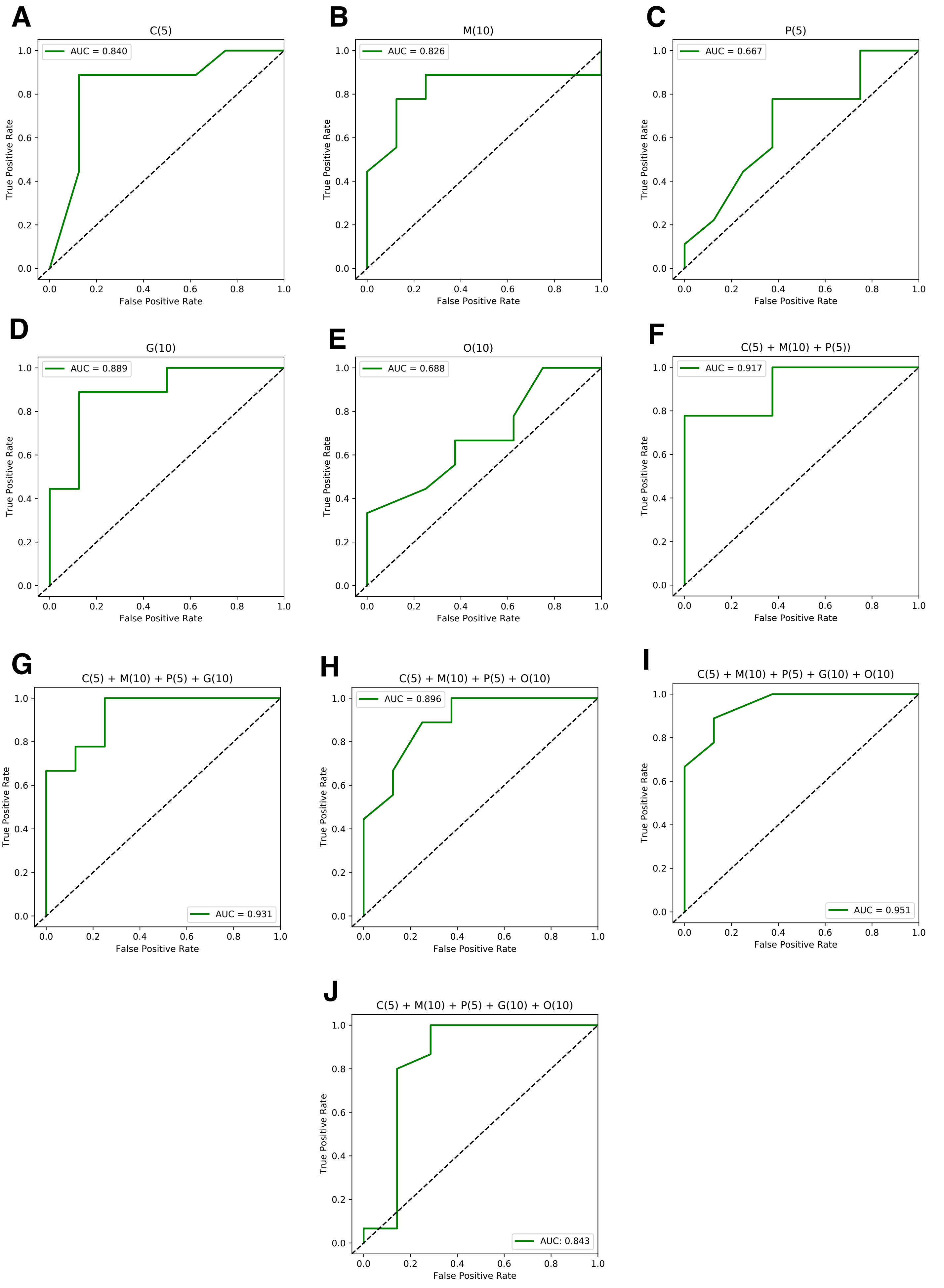
